# Omicron reactive multi protein specific CD4 T cells defines cellular immune response induced by inactivated virus vaccines

**DOI:** 10.1101/2022.05.25.22275616

**Authors:** Joey Ming Er Lim, Shou Kit Hang, Smrithi Hariharaputran, Adeline Chia, Nicole Tan, Eng Sing Lee, Edwin Chng, Poh Lian Lim, Barnaby Young, David Chien Lye, Nina Le Bert, Antonio Bertoletti, Anthony T Tan

## Abstract

Unlike mRNA vaccines based only on the Spike protein, inactivated SARS-CoV-2 vaccines should induce a diversified T cell response recognizing distinct structural proteins. Here we performed a comparative analysis of SARS-CoV-2 specific T cells in healthy individuals following vaccination with inactivated SARS-CoV-2 or mRNA vaccines. Relative to Spike mRNA vaccination, inactivated vaccines elicited a lower magnitude of Spike-specific T cells, but the combined Membrane, Nucleoprotein and Spike-specific T cell response was quantitatively comparable to the sole Spike T cell response induced by mRNA vaccines, and they efficiently tolerate the mutations characterizing the Omicron lineage. However, this multi-protein specific T cell response was not mediated by a coordinated CD4 and CD8 T cell expansion but by selected priming of CD4 T cells. These findings can help in defining the role of CD4 and CD8 T cells in the efficacy of the different vaccines to control severe COVID-19 after Omicron infection.

## Introduction

The availability of vaccines against SARS-CoV-2 have altered the landscape of the COVID-19 pandemic and allowed the virus to persist among vaccinated populations. Globally, numerous SARS-CoV-2 vaccines have been developed, clinically trialled and administered till date^1^ and vaccines based on inactivated virus (Coronavac and Sinopharm) have been utilized in almost half of the 7.3 billion doses that have been delivered to people worldwide before the end of 2021^2^. Despite this staggering number, a detailed analysis of cellular immune response elicited by inactivated vaccines in comparison to mRNA or adenoviral based vaccines is still lacking. The reasons for the lack of a comparative analysis of the components of the adaptive immunity responsible for killing virus-infected cells (virus-specific CD8 T cells), aiding the production of high affinity antibodies (T follicular helper T cells) and sustaining CD8 T cell function (Th1) are different^3^. Inactivated vaccines were utilized largely in China where Western vaccines based on mRNA or adenoviral vectors were not available for a parallel comparison, or in less wealthy nations that often lack research infrastructure to perform the complex characterization of T cell response. Furthermore, the evaluation of the efficacy of different vaccines were performed before the emergence of the antibody escaping Omicron variant^1^, where clinical and virological parameters, like the protection from infection or from symptomatic disease, were associated with the quantity of neutralizing antibodies^4^. These comparative studies showed that the quantity of neutralising antibodies stimulated by inactivated vaccines were ∼10 times lower than that induced by Spike mRNA vaccines and that significant waning of the antibodies occur approximately 3 months post-vaccination^5–9^, faster than the decline of antibodies observed with mRNA vaccines that appears to persist at high levels for at least 6 months^10–16^.

These antibody response comparisons are however now obsolete since the evaluation of vaccination efficacy against Omicron is now directed more towards understanding the ability of vaccines to protect from disease and not infection^17^. Here, T cells are likely to play a more important role in light of their ability to tolerate most of the amino acid variations present in Omicron and their ability to target virus infected cells^10,18,19^. In this regard, real world vaccine efficacy data showed that individuals vaccinated with inactivated virus developed in general milder disease than non-vaccinated individuals^20–23^. However, a direct comparative analysis of individuals vaccinated with inactivated SARS-CoV-2 and mRNA vaccines showed relatively lowered protection from infection and severe disease against Delta lineage in the former^16^, in line with a meta-analysis comparing the clinical efficacy of multiple COVID-19 vaccines^24^. These data suggest that inactivated vaccines might actually induce not only a weaker humoral response towards Spike but also an impaired level of T cell response which was however not experimentally supported. Analysis of SARS-CoV-2 specific T cells in individuals vaccinated with inactivated virus demonstrated the induction of T cells specific for Spike and other structural proteins (NP and Membrane)^25,26^ and a magnitude of vaccine-induced CD8 T cells that were superior to the Spike-specific CD8 T cells induced by mRNA vaccines^26^. Such strong induction of CD8 T cell response was perplexing since the antigen processing and presentation pathway associated with an exogenous protein antigen like inactivated SARS-CoV-2 vaccine is expected to induce primarily a CD4 T cell response^27^. Even though adjuvants can increase cross-presentation of viral antigen to CD8 T cells mediated by a specialized population of dendritic cells^28^, such processes should be less efficient than the direct presentation of viral antigen by MHC-class I occurring in vaccine preparations where antigen is endogenously synthesized within the cells, like in the case of mRNA based vaccines.

We therefore longitudinally analysed the vaccine-specific T cells induced in healthy individuals after SARS-CoV-2 vaccination with inactivated or mRNA vaccines and characterized their ability to recognize multiple proteins and the involvement of CD4 and CD8 T cells in such response. In addition, some of the inactivated SARS-CoV-2 vaccinees received a homologous booster within the study period of 6 months. The effects of boosting on the vaccine-induced T cell response was also analysed together with the evaluation of the effects of mutations present in the Omicron VOC on the vaccine-induced T cell response against multiple viral proteins.

## Results

### Magnitude of Spike-specific T cell response following vaccination with inactivated SARS-CoV-2 vaccines

Utilizing a pool of peptides covering the major immunodominant regions of the Spike protein (SpG pool) in a whole blood cytokine release assay (CRA) that we have previously developed and validated in mRNA vaccinated or infected individuals^29^, we longitudinally analysed the magnitude of Spike-specific T cell response following vaccination with inactivated SARS-CoV-2 vaccines in two cohorts (Figure 1A; Table 1). The first cohort (Inactivated cohort) are healthy individuals who received two doses of BBIBP inactivated vaccine (Sinopharm) given 21 days apart (n=30). The second cohort (Heterologous cohort) are healthy individuals who first received a single dose of Spike mRNA vaccine (Pfizer-BioNTech or Moderna) and developed adverse events that precludes the administration of the second dose of mRNA vaccine. After the normalisation of the adverse events (1-6 months after occurrence of adverse events), these individuals received two doses of CoronaVac inactivated vaccine (Sinovac) given 21 days apart (n=20). A reference cohort (mRNA cohort) of healthy individuals who received two doses of Spike mRNA vaccine (Pfizer-BioNTech or Moderna) were also analysed with the whole blood CRA (n=79).

**Figure 1.**
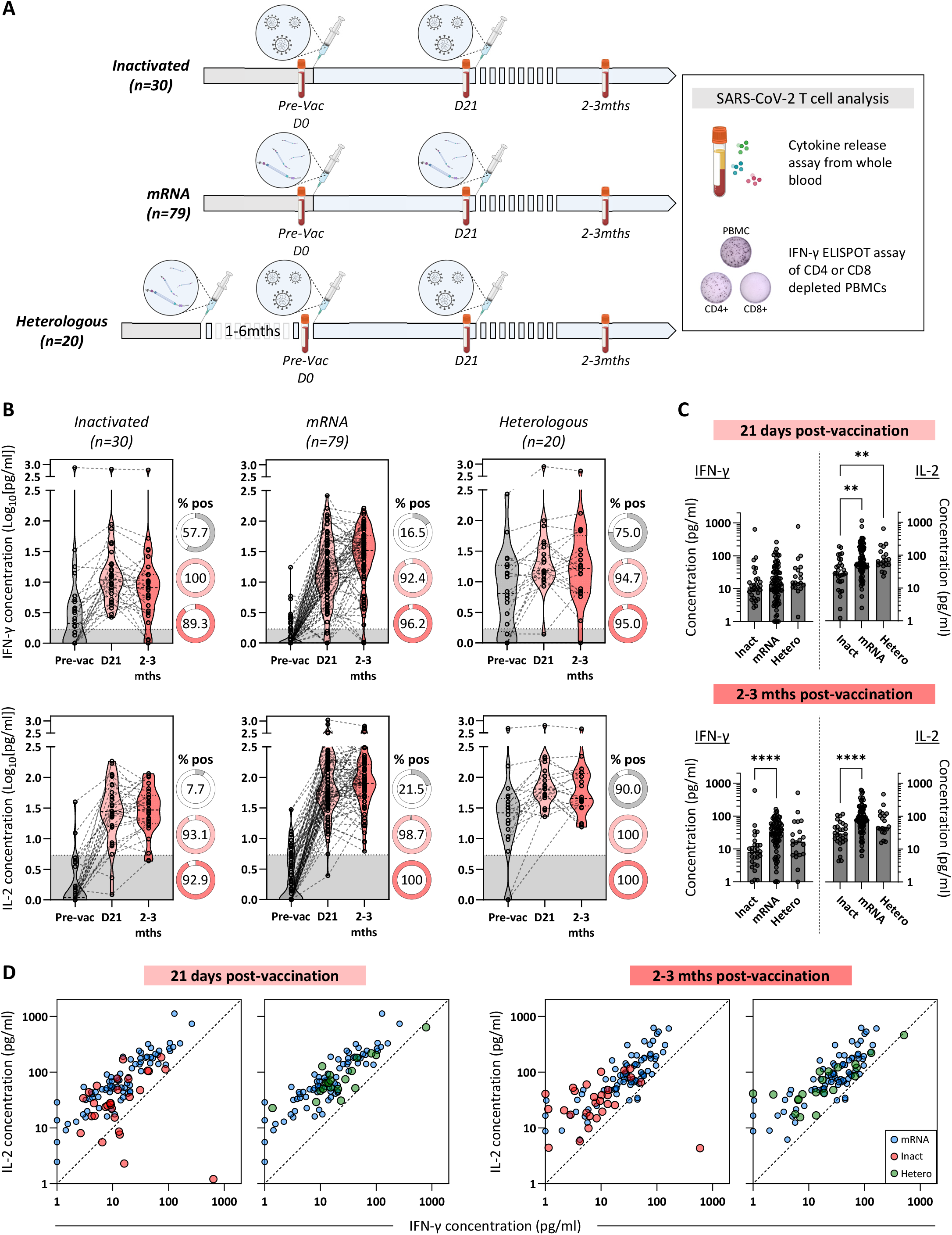
Lower quantities of Spike-specific T cells were induced following vaccination with inactivated SARS-CoV-2 compared to Spike mRNA vaccines. A) Schematic showing the different groups of vaccinated individuals studied and the time points where SARS-CoV-2 specific T cells were analysed. Healthy individuals were given two infusions of either inactivated SARS-CoV-2 vaccines (Inactivated: n=30) or Spike mRNA vaccines (mRNA: n=79) spaced 21 days apart. In the third group (Heterologous: n=20), healthy individuals were first given a single infusion of Spike mRNA vaccine, followed by two infusions of inactivated SARS-CoV-2 vaccine spaced 21 days apart due to the development of significant adverse events following mRNA vaccination. Blood collected at the indicated time points were used to quantify SARS-CoV-2 specific T cells. B) Longitudinal IFN-γ and IL-2 concentrations detected through whole blood CRA after stimulation with SpG peptide pool. Grey shaded areas denote the positivity cut-off for the measured cytokines. Pie charts shows the proportion of individuals with detectable SpG-reactive T cells at the different time points. C) Cross-sectional comparison of median IFN-γ and IL-2 concentrations detected through whole blood CRA after stimulation with SpG peptide pool. Differences were analysed with Kruskal-Wallis test and multiple comparisons were adjusted with Dunn’s multiple comparison test. Only significant adjusted P values are shown (****: P≤0.0001; ***: P≤0.001; **: P≤0.01; *: P≤0.05). D) Dot plots shows the IFN-γ and IL-2 concentrations detected through whole blood CRA after stimulation with SpG peptide pool at the indicated time points. Each dot denotes a single individual. Individuals in the Inactivated cohort (red dots) and the Heterologous cohort (green dots) were overlaid on the vaccinees from the mRNA cohort (blue dots). A dashed line of identity was added for reference.

**Table 1.** Demographics of vaccinees from the 3 study cohorts.

|  | **Heterologous** | **Inactivated** | **mRNA** |
| --- | --- | --- | --- |
| **Characteristic** | One dose of mRNA vaccine (BNT162b2 or mRNA-1273) followed by two doses of CoronaVac | Two doses of BBIBP-Corv, 21 days apart | Two doses of BNT162b2, 21 days apart |
| **N** | 20 | 30 | 79 |
| **Age** | Mean (range): 47.54 (22-69) | Mean (range): 48.7 (22-74) | Mean (range): 38.1 (22-60) |
| **Sex** | F: 14<br>M: 6 | F: 16<br>M: 14 | F: 55<br>M: 24 |
| **Race** | Chinese: 18<br>Malay: 2 | Chinese: 26<br>Indian: 1<br>Malay: 1<br>Others: 2 | Chinese: 42<br>Indian: 7<br>Malay: 10<br>Others: 20 |

A single dose of inactivated vaccine induced an increase in Spike-induced IFN-γ and/or IL-2 in all individuals of the Inactivated cohort 21 days post-vaccination (Figure 1B). In the Heterologous cohort, Spike induced cytokine responses were already present before receiving inactivated vaccines due to the prior dose of Spike mRNA vaccine, and the response remained high after inactivated SARS-CoV-2 vaccination (Figure 1B). The level of cytokines induced by the Spike peptide pool remained stable for up to 2-3 months post-vaccination (1-2 months after receiving the second dose) in both cohorts (Figure 1B). Even with the initial priming from mRNA vaccines prior to inactivated SARS-CoV-2 boosting, the peak magnitude of the Spike-induced cytokines detected in the Heterologous cohort do not differ from the reference mRNA cohort (Figure 1B and 1C).

While the response kinetics appear similar between the Inactivated and mRNA cohorts, with peaks of Spike-induced IFN-γ and IL-2 observed 21 days post-vaccination, the magnitude of the Spike-induced response was lower in individuals who received inactivated vaccines than compared to those who had Spike mRNA vaccination (Figure 1B and 1C). This difference was particularly significant 2-3 months post-vaccination (Figure 1C). Even in the Heterologous cohort where individuals were primed with Spike mRNA vaccine before boosting with inactivated vaccines, the magnitude of the Spike-induced T cell cytokines was lower but did not reach statistical significance (Figure 1C). Analysis of the quantities of IFN-γ and IL-2 secreted after SpG peptide pool stimulation did not show significant differences between all 3 cohorts at 21 days or 2-3 months post-vaccination (Figure 1D).

These results shows that inactivated SARS-CoV-2 vaccines induced a quantitatively lower Spike-specific T cell response with minimal differences in the balance of IFN-γ and IL-2 secreted in comparison to mRNA vaccines. Consistent with what was observed previously^29^, quantifying the Spike T cell response through IL-2 secretion also appears more robust with less variability than by IFN-γ secretion (Figure 1).

### Immunodominance of Spike-specific T cells induced by inactivated SARS-CoV-2 vaccines

To understand the breadth of the Spike-specific T cell response induced by inactivated SARS-CoV-2 vaccines, we also performed the whole blood CRA assay using 7 overlapping peptide pools that spans the entire Spike protein on blood collected 2-3 months after receiving inactivated or mRNA vaccines. A schematic representation of the localization of peptide pools 1 to 7 in relation to the S1 (N-terminal), RBD and S2 (C-terminal) regions of Spike is displayed in Figure 2A. Randomly selected individuals from the Inactivated (n=5) and Heterologous (n=8) cohorts were analysed and we detected Spike T cell responses in all individuals (Figure 2B). Almost all of the tested individuals had dominant Spike-specific T cell response targeting the S2 chain of the Spike protein (Spike pool 5-7) (Figure 2B and 2C), with the strongest response targeting pool 6 that covers amino acid 886-1085 of the Spike protein. There were no observable differences between the cohorts and this immunodominance pattern is similar to that observed in a previous study on mRNA vaccinated individuals (Figure 2B, mRNA n=6) where majority of the Spike-specific T cells were targeting epitopes present in the S2 chain of Spike^29^.

**Figure 2.**
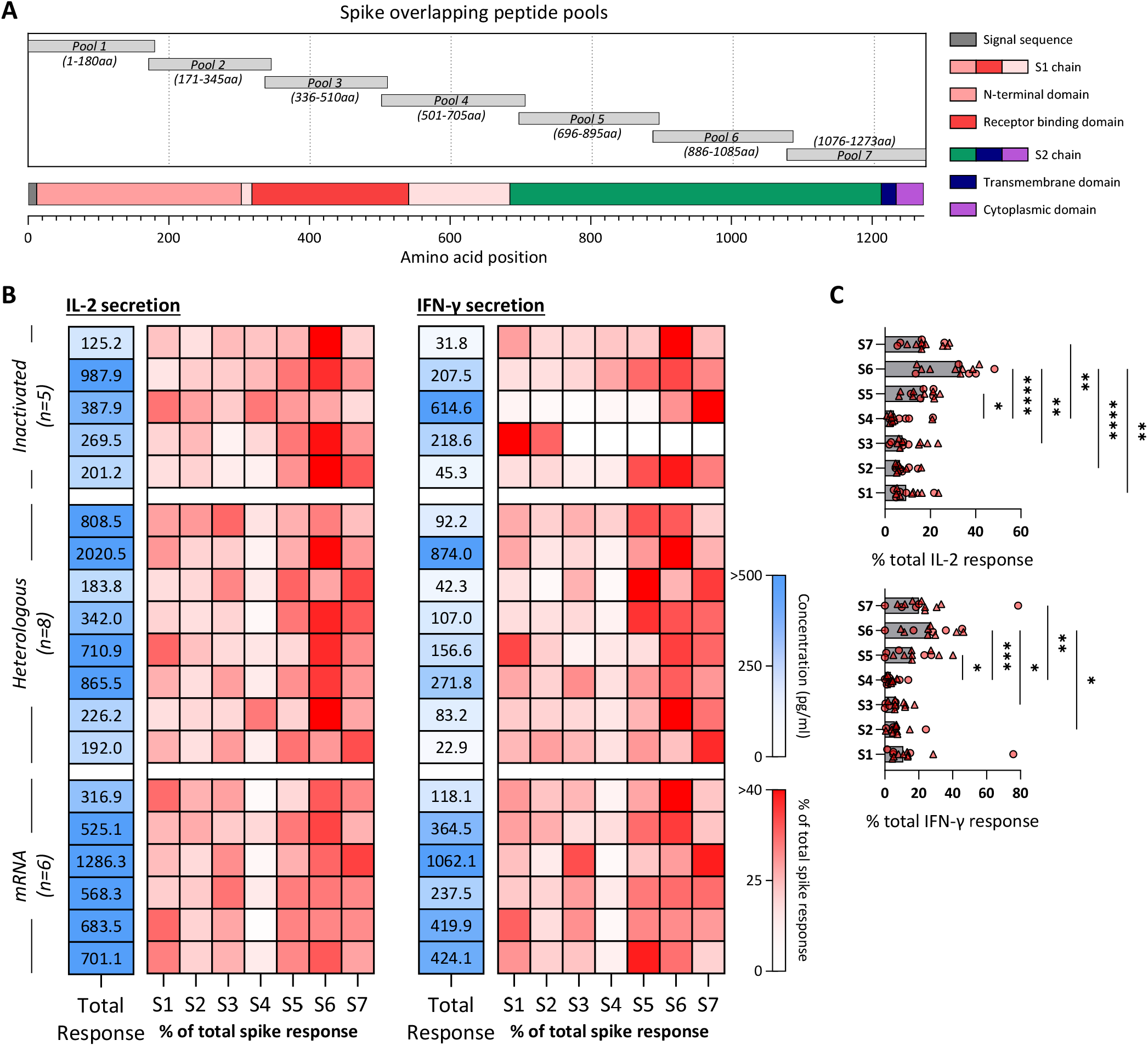
Inactivated SARS-CoV-2 vaccine-induced Spike-specific T cells targets the S2 chain of the spike protein. A) Schematic representation of the 7 Spike-specific peptide pools containing 15-mer overlapping peptides spanning the entire Spike protein. Pools 1-4 contain peptides from the signal peptide and the S1 chain while pools 5-6 encompass the S2 chain together with the transmembrane and cytoplasmic domains. B) The amount of IFN-γ and IL-2 secreted in the whole blood CRA in response to overlapping peptides covering the entire Spike protein (total Spike-specific T cell response) of all vaccinees 2-3 months after receiving inactivated (Inactivated: n=5; Heterologous: n=8) or mRNA vaccines (mRNA: n=6) are shown in the blue heatmap. The red heatmap denotes the proportion of Spike-specific T cell responses of each vaccinee targeting the different parts of the Spike protein. C) Bars shows the median proportion of Spike-specific T cell responses targeting the different parts of the Spike protein as assayed through IFN-γ and IL-2 whole blood CRA in the vaccinees from the Inactivated (circles) and Heterologous (triangles) cohorts. Differences were analysed with Kruskal-Wallis test and multiple comparisons were adjusted with Dunn’s multiple comparison test. Only significant adjusted P values are shown (****: P≤0.0001; ***: P≤0.001; **: P≤0.01; *: P≤0.05).

### Inactivated vaccine-induced T cell responses targeting multiple viral structural proteins

The breadth of inactivated SARS-CoV-2 vaccine-induced T cell response should extend beyond the Spike protein since the inactivated vaccine contains other structural proteins in addition to Spike^30,31^. Similar to the analysis of Spike-specific T cell response, we used overlapping peptides covering the entire Membrane and Nucleoprotein in a whole blood CRA. We observed induction of Membrane-(Figure 3A) and Nucleoprotein-specific (Figure 3B) T cell response in both cohorts. Over 90% of the individuals in both cohorts had detectable Membrane- and Nucleoprotein-specific T cell response 21 days after vaccination, and the response persisted for at least 2-3 months post-vaccination (Figure 3A and 3B). Secretion of IL-2 was again more robust with less variability among the individuals. Importantly, the magnitude of the Spike T cell response was positively correlated with both the Membrane and Nucleoprotein T cell response (Figure 3C), demonstrating that these responses were indeed induced and associated with inactivated SARS-CoV-2 vaccination. Comparative analysis of the cytokine quantity induced by Spike, Membrane and Nucleoprotein peptide pools 2-3 months post-vaccination, showed that the majority of the vaccine induced T cell response were targeting the Spike protein, followed by Nucleoprotein and Membrane, a hierarchy proportional to the size of the respective proteins (Figure 3D).

**Figure 3.**
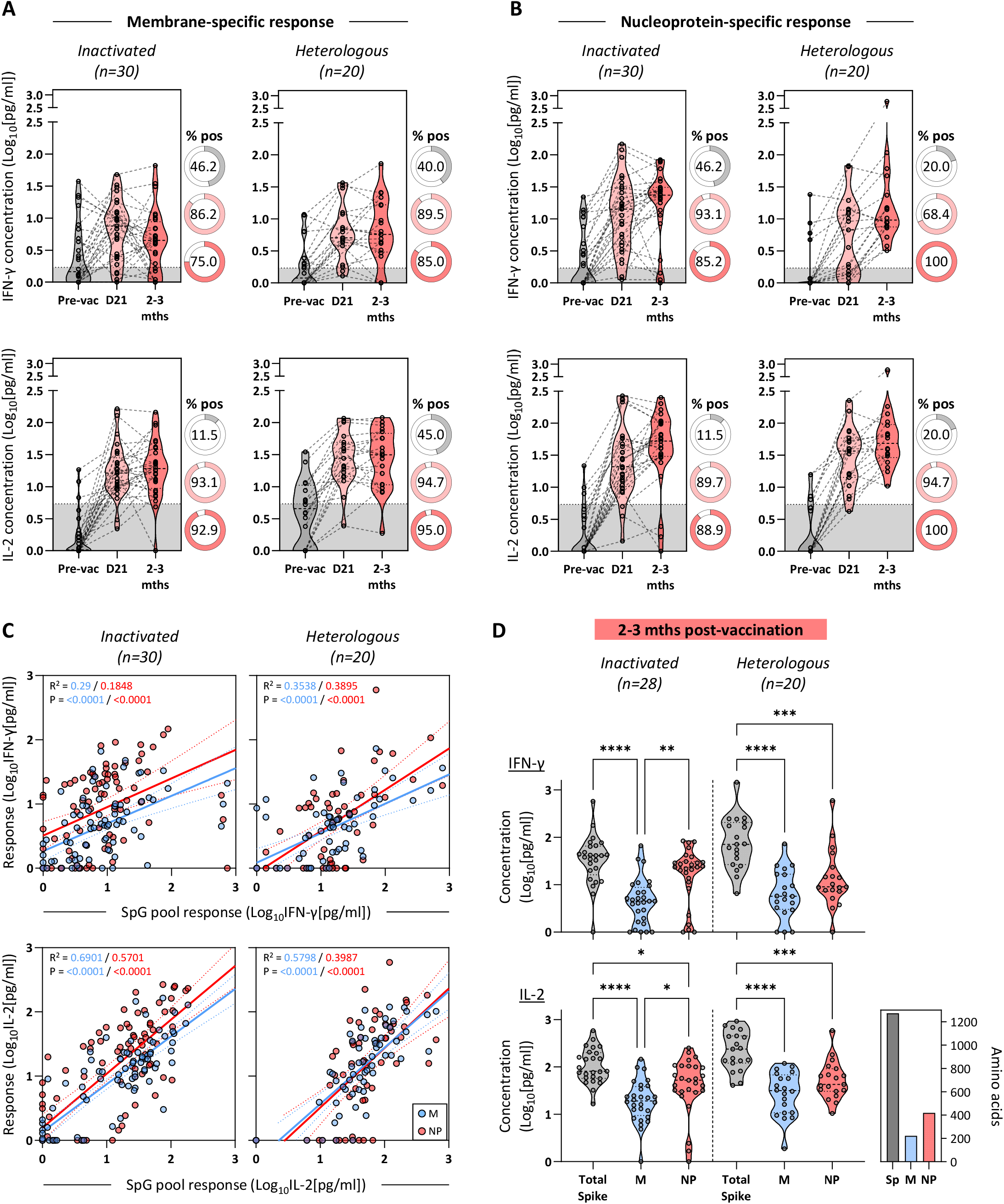
T cell responses targeting Membrane and Nucleoprotein were expanded following vaccination with inactivated SARS-CoV-2. Longitudinal IFN-γ and IL-2 concentrations detected through whole blood CRA of all vaccinees receiving inactivated SARS-CoV-2 vaccines (Inactivated: n=30; Heterologous: n=20) after stimulation with overlapping peptides covering the entire Membrane (A) and Nucleoprotein (B). Grey shaded areas denote the positivity cut-off for the measured cytokines. Pie charts shows the proportion of individuals with detectable M- or NP-reactive T cells at the different time points. C) Linear regression analysis of the T cell response against SpG peptide pool and the Membrane (blue) or Nucleoprotein (red) as evaluated by the quantification of IFN-γ (top) or IL-2 (bottom) in peptide stimulated whole blood from vaccinees (Inactivated: n=30; Heterologous: n=20) at all analysed time points. Dotted lines denote the 95% confidence interval. D) Total Spike-, Membrane- and Nucleoprotein-specific T cell response detected through whole blood CRA (IFN-γ: top or IL-2: bottom) of all vaccinees 2-3 months after receiving inactivated vaccines (Inactivated: n=28; Heterologous: n=20). Bar graph insert shows the size (amino acid) of the indicated proteins. Differences were analysed with Kruskal-Wallis test and multiple comparisons were adjusted with Dunn’s multiple comparison test. Only significant adjusted P values are shown (****: P≤0.0001; ***: P≤0.001; **: P≤0.01; *: P≤0.05).

We also measured the global level of T cell response induced by inactivated SARS-CoV-2 and Spike mRNA vaccines. The amount of IFN-γ or IL-2 secreted after stimulation with SpG, Membrane and Nucleoprotein peptide pools in both inactivated (n=27) and Heterologous (n=18) vaccinated cohorts were comparable to the one detected by SpG only in mRNA vaccine recipients (n=79) (Figure 4).

**Figure 4.**
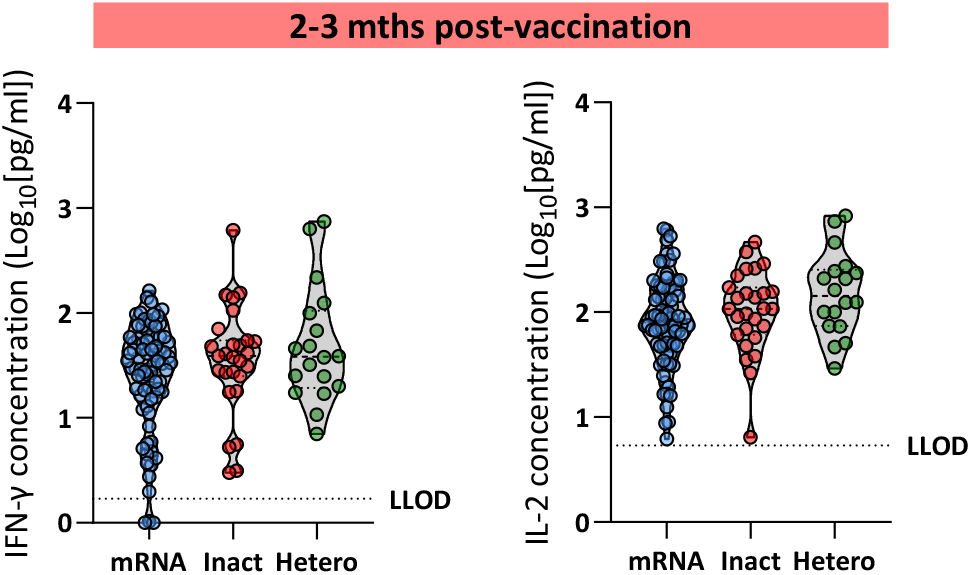
Total T cell response induced by inactivated SARS-CoV-2 and mRNA vaccines are comparable. Total vaccine-specific T cell response (Spike only for mRNA vaccine; Spike, Membrane and Nucleoprotein for inactivated vaccines) detected through whole blood CRA (IFN-γ: left or IL-2: right) of vaccinees 2-3 months after receiving the respective vaccines (mRNA: n=79; Inactivated: n=27; Heterologous: n=18). Spike-specific T cell response were evaluated by stimulation with SpG peptide pool, while overlapping peptide megapools covering the entire Membrane or Nucleoprotein were used to quantify the respective protein-specific T cell response. Dotted lines denote the lower limit of quantification (LLOD) for each of the measured cytokine. Differences were analysed with Kruskal-Wallis test and multiple comparisons were adjusted with Dunn’s multiple comparison test. Only significant adjusted P values are shown (****: P≤0.0001; ***: P≤0.001; **: P≤0.01; *: P≤0.05).

### Phenotype of vaccine-induced T cell responses against multiple viral proteins

We next sought to determine whether the inactivated vaccine-induced T cell response were mediated by helper CD4+ or cytotoxic CD8+ T cells. Two different methods were used: a classical analysis of upregulation of T cell activation markers (AIM) on the CD4 (CD25+ OX40+ 41BB+) and CD8 (CD69+ 41BB+) T cell subsets after PBMC stimulation with peptide megapools, or a depletion approach where CD8 and CD4 T cells were removed from the PBMC before stimulation with the peptide indicated megapools in an IFN-γ ELISPOT assay (Figure 5A).

**Figure 5.**
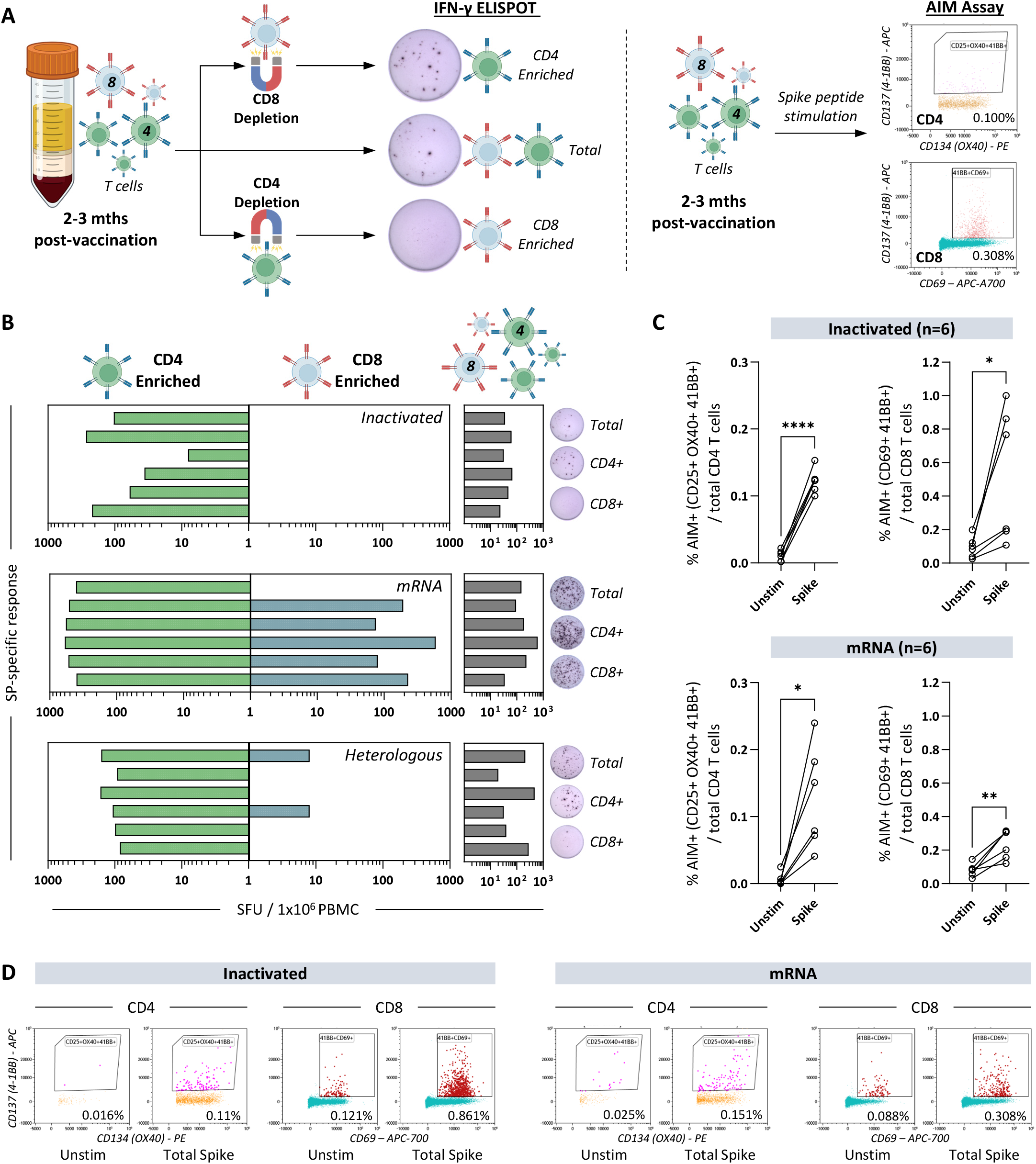
Inactivated SARS-CoV-2 vaccines stimulates primarily CD4 T-cell responses. A) Schematic illustrates the analysis of SARS-CoV-2 specific CD4 and CD8 T cells by IFN-γ ELISPOT and by the detection of activation markers on CD4 (CD25+ OX40+ 41BB+) and CD8 (CD69+ 41BB+) T cells. PBMCs collected from the 3 cohorts of vaccinees 2-3 months after vaccination (Inactivated: n=6; mRNA: n=6; Heterologous: n=6) were either depleted of CD8 or CD4 T cells through negative selection before stimulation with overlapping peptides covering the entire Spike for ELISPOT. Total PBMCs were also analysed as a control. For the AIM assay, total PBMCs were stimulated with overlapping peptides covering the entire Spike protein for 24 hours before flow cytometry analysis. B) Bars denote the IFN-γ SFU quantified for each vaccinee after stimulating CD4-enriched (green), CD8 enriched (blue) or total PBMCs (grey) with Spike overlapping peptides. Representative ELISPOT well images of the respective peptide stimulated cell populations are shown. C) Flow cytometry analysis of activation marker expression on CD4 and CD8 T cells from individuals in the Inactivated (n=6) and mRNA (n=6) cohort before and after stimulation with overlapping peptides covering the entire Spike protein. Differences were analysed with paired t-test and only significant P values are shown (****: P≤0.0001; ***: P≤0.001; **: P≤0.01; *: P≤0.05). D) Flow cytometry dot plots of CD4 and CD8 T cells from representative individuals before and after Spike peptide stimulation are shown together with the corresponding AIM+ frequencies among the total CD4 or CD8 T cells.

The two methods yielded completely different results in terms of the composition of the peptide pools responsive T cells. A significant upregulation of AIM on both CD4 and CD8 T cells upon Spike stimulation was detected in all tested individuals from both Inactivated and mRNA cohorts (Figure 5C and D). In contrast, using the depletion approach, we observed a clear CD4 centric Spike T cell response induced by inactivated vaccines (Figure 5B). Spike-specific CD8 T cells were only observed at very low quantities in 2 out of 12 individuals vaccinated with inactivated SARS-CoV-2, in contrast to the Spike-specific CD4 T cells that were present in all individuals tested (Figure 5B). In addition, T cells specific for Nucleoprotein and Membrane were also CD4 centric with no detectable response in the CD8 enriched samples (Supplementary Figure 3). On the other hand, both CD4 and CD8 Spike-specific T cells were induced by mRNA vaccines (Figure 5B).

### Effects of inactivated SARS-CoV-2 vaccine booster (third dose) on the T cell response

Studies that follows individuals who received primary vaccination (2 doses) with inactivated SARS-CoV-2 vaccines have reported a progressive waning of neutralizing antibody at 3 months with titres almost reaching pre-vaccination levels at ∼6 months^6–8^. This has prompted policy changes in Singapore to recommend the inclusion of a third dose of inactivated SARS-CoV-2 into the primary vaccination series 3 months after the administration of the second dose. While the additional dose of inactivated SARS-CoV-2 vaccine have been shown to effectively recall the neutralizing antibody titres that have declined substantially after the second dose^7^, little is known about its effects on the amplification of vaccine-induced T cell responses. As such, we compared the vaccine-induced T cell response before and after the third vaccine dose in the Inactivated and Heterologous cohorts using the whole blood CRA with SpG, Membrane and Nucleoprotein peptide pools.

In the cohorts studied, a total of 18 individuals in the Inactivated cohort and 7 in the Heterologous cohort were given the third vaccine dose 1-3 months and <1 month before blood collection at ∼6-7 months after the first vaccination dose respectively (Figure 6A). 3 individuals in the Inactivated cohort and 11 in the Heterologous cohort have not received the third dose of vaccine at the time of blood collection and they were analysed as controls. None of the vaccinated individuals analyzed had a known history of SARS-CoV-2 infection up until the last blood sample collection at ∼6-7 months from the first vaccination dose.

**Figure 6.**
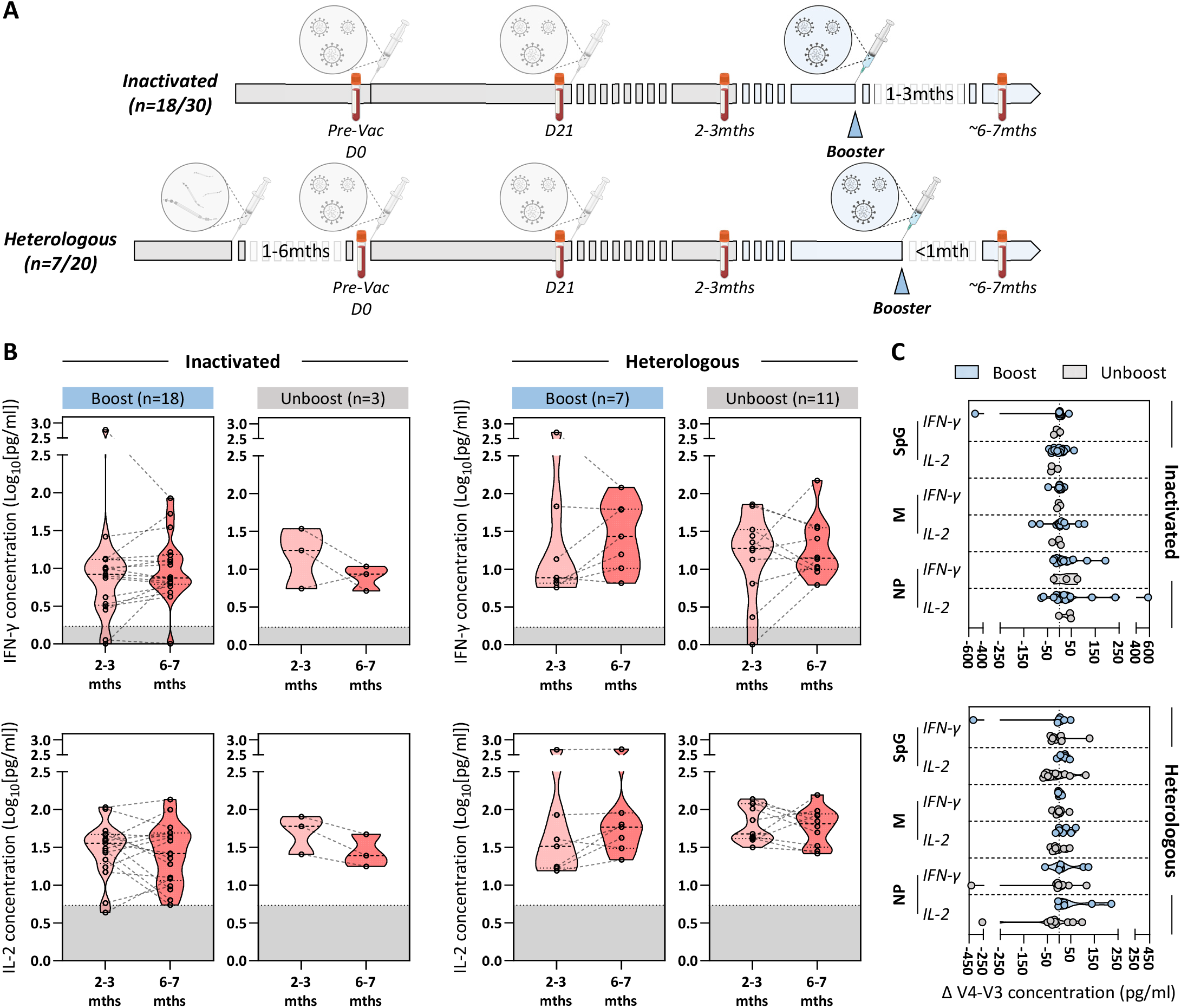
Vaccine induced T cell responses were not boosted upon the third inactivated vaccine dose. A) Schematic showing the time points where a third dose of inactivated vaccine was given to individuals in the Inactivated (n=18/30 boosted) and Heterologous (n=7/20 boosted) cohorts. Blood was collected ∼6-7 months after the first dose of inactivated vaccines, which corresponds to 1-3 months and <1 month after the booster dose for individuals in the Inactivated cohort and Heterologous cohort respectively. SARS-CoV-2 specific T cells after the booster dose were quantified as described previously (Figure 1 and 3) through whole blood CRA after stimulation with SpG, Membrane and Nucleoprotein peptide pools. Individuals who did not received the booster dose were analysed as controls. B) Longitudinal IFN-γ and IL-2 concentrations detected through whole blood CRA after stimulation with SpG peptide pool in individuals before and after receiving the booster dose. Grey shaded areas denote the positivity cut-off for the measured cytokines. C) Graph shows the change in IFN-γ and IL-2 concentrations detected through whole blood CRA after stimulation with SpG, Membrane and Nucleoprotein peptide pools before and after the booster dose (V4-V3 concentrations) in individuals from the Inactivated and Heterologous cohorts. Differences were analysed with Kruskal-Wallis test and multiple comparisons were adjusted with Dunn’s multiple comparison test. Only significant adjusted P values are shown (****: P≤0.0001; ***: P≤0.001; **: P≤0.01; *: P≤0.05).

From the whole blood CRA, we did not observe an appreciable change in the amount of secreted IFN-γ and IL-2 after SpG peptide pool stimulation of whole blood samples collected before (∼1-2 months after the 2^nd^ dose) and after (either 1-3 months or <1 month after the 3^rd^ dose) the third dose of inactivated SARS-CoV-2 vaccine in both Inactivated and Heterologous cohorts (Figure 6B). The response towards Membrane and Nucleoprotein also remained unchanged before and after the third vaccination dose (Figure 6C). Importantly, the magnitude of the whole blood CRA response against all peptide pools tested did not differ between the individuals in either cohorts who received the third inactivated vaccine dose and the controls who did not (Figure 6B and C).

Thus, the third dose of inactivated SARS-CoV-2 vaccine did not effectively induce a significant boost of the vaccine-induced T cell response stimulated by the first two inactivated vaccine doses. Unlike the neutralizing antibody titres that wanes within 3-6 months of the second dose^6–8^, the inactivated vaccine-induced T cell response was maintained for at least 6 months without observable waning (Figure 6B and C).

### Inactivated vaccine-induced T cell responses are largely preserved against the Omicron variant of concern

Given that the Omicron variant of concern (VOC) is the prevailing SARS-CoV-2 lineage circulating globally, we determined whether the inactivated vaccine-induced T cell responses can recognise the Omicron VOC proteins. We designed 3 sets of peptide pools specific for the Spike, Membrane and Nucleoprotein respectively (Supplementary Table 1). Each set consist of a megapool made up of overlapping peptides spanning the entire vaccine-derived protein, a WT pool containing peptides covering the regions affected by mutations present in the Omicron variant of the protein, and a MT pool consisting of peptides from the WT Pool with the amino acid mutations present in the Omicron variant of the protein (Figure 7A). These peptide pools were used in the whole blood CRA using samples collected from individuals in the Inactivated and Heterologous cohorts ∼6-7 months after their first inactivated vaccine dose. Only individuals who received the third inactivated vaccine dose were analysed to ensure that the vaccine-induced T cell response was at its optimum.

**Figure 7.**
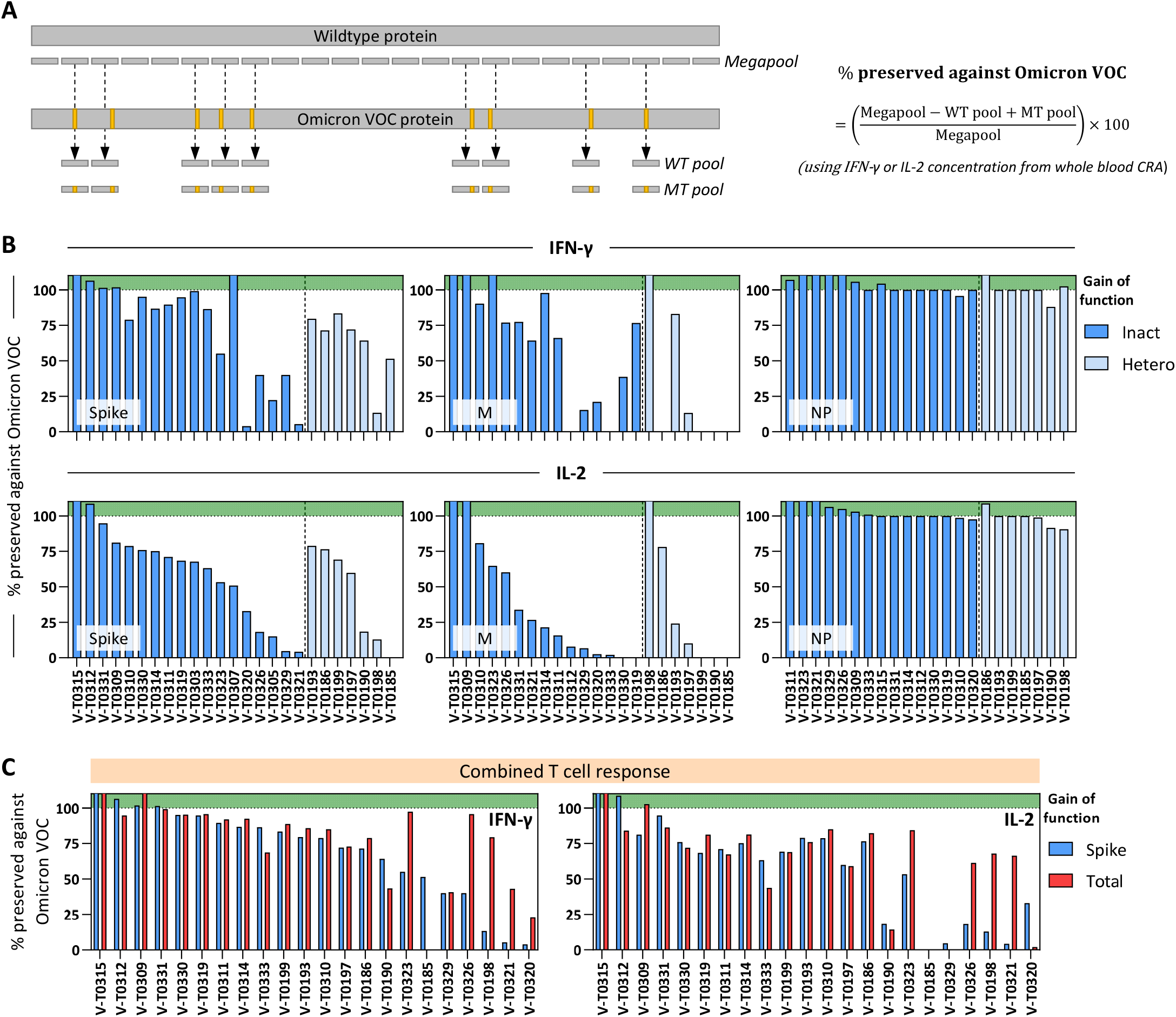
Inactivated SARS-CoV-2 vaccine-induced T cell response against the Omicron variant of concern. A) Schematic showing the concept behind the design of the peptide pools to assess the inactivated SARS-CoV-2 vaccine-induced T cell response against the Spike, Membrane and Nucleoprotein from the Omicron variant of concern. Orange regions refers to amino acid mutations present in the Omicron variant relative to the vaccine-derived SARS-CoV-2. The respective Megapool contains peptides covering the whole wildtype SARS-CoV-2 protein (Spike, Membrane or Nucleoprotein). The respective WT Pool contains peptides, with the wildtype amino acid sequence, covering the regions affected by mutations present in the Omicron variant of the protein. The respective MT Pool contains peptides from the WT Pool with the amino acid mutations present in the Omicron variant. The equation shows how the percentage preservation of the vaccine-induced T cell response against the Spike, Membrane or Nucleoprotein from the Omicron variant of concern was calculated. B) Inactivated SARS-CoV-2 vaccine-induced T cell response of individuals from the Inactivated (n=18; dark blue) and Heterologous (n=7; light blue) cohorts were evaluated by whole blood CRA using the peptide pools described in A. The percentage preservation of the vaccine-induced T cell response against the Spike, Membrane and Nucleoprotein from the Omicron variant of concern were calculated using IFN-γ (top) or IL-2 (bottom) concentrations from whole blood CRA and displayed as bars. Bars that enter the green shaded areas refers to a gain of function response against the Omicron variant of concern. C) The percentage preservation of the Spike (blue) and total inactivated vaccine-induced T cell response (red; Spike, Membrane and Nucleoprotein) against the Omicron variant of concern in vaccinees from both Inactivated and Heterologous cohorts were calculated using IFN-γ (left) or IL-2 (right) concentrations from whole blood CRA and displayed as bars.

We observed a wide heterogeneity in the ability of inactivated vaccine-induced T cell response to recognise the Spike and Membrane proteins from the Omicron VOC, which ranges from the complete abrogation to a gain of function exceeding the response towards the corresponding vaccine-derived proteins (Figure 7B). Based on the amount of IFN-γ secreted in the whole blood CRA, 13/18 individuals from the Inactivated cohort and 6/7 individuals from the Heterologous cohort have at least 50% of their Spike-specific T cell response preserved against the Omicron Spike protein (Figure 7B). For the Membrane-specific T cell response, at least 50% of the response was maintained in 10/18 individuals from the Inactivated cohort and 2/7 individuals in the Heterologous cohort (Figure 7B). Interestingly, almost all of the Nucleoprotein-specific T cell response was preserved with no individual exhibiting lower than 80% of response against the Omicron Nucleoprotein (Figure 7B). This is in stark contrast to the Membrane-specific T cell response even though both proteins have similar number of mutations present in the Omicron VOC (Supplementary Table 1).

Combining the T cell response against multiple proteins, majority of the vaccinees still maintained their response towards the Omicron VOC (Figure 7C). Importantly, in vaccinees where the singular Spike-specific T cell response was significantly inhibited, the combined multi-antigenic T cell response showed better preservation against Omicron (Figure 7C). Similar observations were made when the amount of secreted IL-2 was analysed (Figure 7B and C).

## Discussion

The virological landscape of the COVID-19 pandemic was radically modified by the emergence of new SARS-CoV-2 lineages able to escape the neutralizing ability of antibodies elicited by vaccines based on the Spike protein of the ancestor Wuhan isolates^32,33^. This necessitates a more comprehensive analysis of vaccine immunogenicity that could not only be based on antibody measurements but also requires an evaluation of cellular immunity^17^. T cells are not playing a role in the prevention of infection but can be extremely important in the control of the viral pathogenesis due to their ability to recognize and lyse virus-infected cells and their association with viral control in acutely infected patients^34^. For this reason, we performed a detailed characterization of the cellular immunity specific for different SARS-CoV-2 proteins elicited by inactivated virus vaccines and mRNA vaccines in a population of healthy adult individuals.

Using a whole blood CRA^29^ that measures the ability of T cells to secrete cytokines directly in whole blood after encounter with specific antigens, we observed that the quantity of Spike peptide-stimulated T cell cytokines were lower in vaccinees who received inactivated vaccines in comparison to those who had Spike mRNA vaccines. However, both vaccine preparations induced a Th1 response with similar IFN-γ and IL-2 secretion profile. Both vaccines also elicit a comparable Spike immunodominance hierarchy focused more on the S2 region of Spike. (Figure 1 and 2). On the other hand, inactivated vaccines do not elicit a response only focused on the Spike protein. Significant quantities of Membrane and Nucleoprotein specific T cells were also induced after vaccination that are clearly absent in individuals vaccinated with mRNA vaccines (Figure 3). A comparison of the multi-antigenic vaccine-induced T cell response calculated by quantifying the levels of IFN-γ and IL-2 elicited by inactivated versus mRNA vaccines showed that inactivated vaccines not only elicited a broader T cell immunity but they also induce a quantitatively comparable T cell response against SARS-CoV-2 relative to mRNA vaccines (Figure 4).

The importance of eliciting a multi-antigenic T cell response against different SARS-CoV-2 proteins should not be underestimated since T cell response in individuals who control SARS-CoV-2 with limited or no symptoms possess T cell responses against different epitopes present in different structural and non-structural proteins^34–38^. In addition, in experimental SARS-CoV-2 challenge of non-human primates and mice vaccinated against Nucleoprotein^39,40^ or Envelope and Membrane^41^, the vaccinated animals had less severe pathology and lower viral loads. This protective effect was associated with the rapid recall of antigen-specific T cell responses without robust humoral immune response, showing the protective capacity of T cells specific for other structural proteins of SARS-CoV-2^39–41^. The broad specificity is also potentially beneficial when considering the effects of amino acid mutations present in the different SARS-CoV-2 proteins of known VOCs. While the inactivated SARS-CoV-2 vaccine induced T cell response was largely preserved against the Omicron VOC, there were differences in the responses against different structural proteins (Figure 7). In all vaccinees tested, Nucleoprotein-specific T cell responses were not affected by the mutations present in Omicron, while both Spike and Membrane specific T cell responses were inhibited to various levels (Figure 7). This is likely explained by the density of amino acid mutations present in each protein (Spike: 1 mutation in 40 amino acids; Membrane:1 in 67; Nucleoprotein: 1 in 80). A protein with a low density of amino acid mutation is more likely to contain T cell epitopes that are fully conserved between the vaccine strain and Omicron. Importantly, in vaccinees where their Spike-specific T cell response was significantly inhibited by the mutations present in Omicron, their combined vaccine-induced T cell response (Spike, Membrane and Nucleoprotein) was better preserved against Omicron (Figure 7C). The presence of multi-protein specific T cells in individuals vaccinated with inactivated virus can therefore provide them with a population of memory T cells more likely to tolerate the frequently found amino acid substitutions in Spike which can partially suppress the Spike-specific T cell response elicited by current Spike mRNA vaccines^42–48^. While the ability of Omicron to escape the full repertoire of Spike-specific T cells induced by mRNA vaccines occurs only in a minority of individuals (10-15%), immune escape can be substantial in particular for Spike-specific CD8 T cells^48^.

However, the potential advantages of the heterogenous specificity of T cells generated by inactivated SARS-CoV-2 vaccination needs to be balanced by our observation that inactivated vaccines do not elicit any CD8 T cell response against any viral proteins. The robust vaccine-induced T cell response was exclusively mediated in inactivated vaccine recipients by CD4 T cells. In contrast, individuals vaccinated with mRNA and non-replicating adenoviral vectored Spike vaccines are capable of inducing both CD4 and CD8 T cell responses as shown previously^10,49–51^, and by our own data. Our results are in contrast with previous observations by other groups that showed the ability of inactivated vaccines to induce both CD4 and CD8 T cell response but are however in line with other data of T cell response observed in other inactivated viral vaccines^52,53^. The difference appears to be due to the different method of analysis utilized by us and by the group in Hong-Kong^26^. The AIM assay utilized by them and by many groups worldwide (including us^29,42,54^) is a powerful cellular technique that can define the phenotype of peptide responsive T cells. It is equally well known that robust cytokine response of peptide responsive T cells can also activate other T cells in the vicinity in a bystander fashion without T cell receptor/epitope engagement^55–57^. From our comparative analysis of results obtained by AIM and by depleting CD4 and CD8 T cells, it seems clear that a robust activation of CD8 T cells was observed in inactivated vaccine recipients only with the AIM assay, showing that the results derived from the AIM assay can be confounded, at least in vaccine recipients by robust bystander activation.

The combined impact of the multi-protein specific T cell response and its association with a deficiency of CD8 T cell induction on the protection from disease development is difficult to be measured. In SARS-CoV-2 convalescent rhesus macaques with sub-protective antibody titres, depletion of CD8 T cells partially abrogated the protective efficacy of natural immunity against viral challenge, showing the anti-SARS-CoV-2 effects mediated directly by CD8 T cells^58^. At the same time, the induction of CD4 T cells specific for the nucleoprotein of SARS-CoV in the nasal cavity of mice protected the animal from lethal disease after infection with different Coronaviruses^59^. This protective effect was mediated through the secretion of IFN-γ and importantly, through the subsequent recruitment of virus-specific CD8 T cells^59^. Similarly, memory CD4 T cells also directly and indirectly mediate protective effects in Influenza A virus infected mice^3^. Clinical analysis of the efficacy of inactivated vaccines in different populations before the emergence of Omicron have shown that inactivated virus vaccines provided protection against the development of severe COVID-19^20–23^ but with lower rates than the one provided by mRNA vaccines. On the contrary, recent data from Hong-Kong measuring the efficacy against mild and severe COVID-19 development in healthy adults infected with Omicron showed similar efficacy of the two vaccine preparations after three doses^60^. Perhaps the lack of the coordinated activation of CD4 and CD8 T cells observed in inactivated virus vaccine recipients infected with Wuhan or Delta strains of SARS-CoV-2 was then compensated by the protein multi-specificity that better tolerate the mutations present in Omicron.

Finally, our work showed that boosting with the third dose of inactivated SARS-CoV-2 vaccine 3 months after the second dose did not modify the Spike, Membrane or Nucleoprotein T cell response (Figure 5). The magnitude of the T cell response against the 3 different proteins were even comparable to individuals who completed their primary vaccination 6 months before and did not receive their booster vaccines within the study period. Our data are in line with what was observed in a phase 2 placebo-controlled trial of inactivated SARS-CoV-2 vaccine: the third dose of inactivated vaccine 2 months after completion of the primary vaccination course (2 doses) only modestly increased the neutralising antibody levels^7^. It is plausible that the ineffective boosting is due to the neutralisation of the administered inactivated SARS-CoV-2 by the antibodies generated from the primary vaccination course leading to a decline in antigen availability to stimulate the immune system. This hypothesis was supported by the observation in the same phase 2 clinical trial where the neutralising antibody titres were significantly boosted and were 3-4 fold higher when the administration of the third vaccine dose was delayed till 8 months (instead of 2 months) after completion of the primary vaccination^7^. It was also observed that humoral and T cell response were both robustly induced when the booster inactivated SARS-CoV-2 vaccine was given 5 months after the primary vaccination^61^. These results certainly calls for the future evaluation of the immunogenicity of inactivated SARS-CoV-2 vaccine in a setting where interference from existing virus-specific immune responses can occur.

In conclusion, we present here a detailed functional and quantitative evaluation of the T cell response induced by inactivated virus vaccines in comparison to mRNA vaccines. We show that in sharp contrast to the clear inferiority of the humoral immunogenicity, inactivated vaccines elicit a T cell response of comparable magnitude and superior breath, relative to the mRNA vaccines, that persisted for at least 6 months without need of further boosting. The ability to recognize different SARS-CoV-2 proteins, in particular Membrane and NP, allowed the T cell response induced by inactivated vaccines to better tolerate the mutations present in Omicron compared to the Spike-focused mRNA vaccine induced T cells. However, the robust SARS-CoV-2 T cell response was exclusively mediated by CD4 T cells while CD8 T cell were not induced. Recent clinical data of vaccine efficacy in a large population suggested that the theoretical defect caused by the biased induction of a single component of cellular immunity by the inactivated vaccine could be balanced by the superior ability to tolerate amino acid mutations present in Omicron. Such data will have to be analysed in a large clinical study where T cell responses will be measured and will help to clarify the impact of virus-specific CD4 or CD8 T cells in SARS-CoV-2 pathogenesis.

## Data Availability

All data produced in the present work are contained in the manuscript

## Acknowledgments

We would like to thank all clinical and nursing staff who provided care for the patients; all clinical trial coordinators and staff for their invaluable assistance in coordinating patient recruitment.

## Author contributions

A.T. Tan, N. Le Bert and A. Bertoletti designed the experiments; J.M.E. Lim, S.K. Hang, S. Hariharaputran, A. Chia, and N. Tan performed the experiments; A.T. Tan, N. Le Bert and A. Bertoletti analyzed and interpreted all the data; A.T. Tan prepared the figures and wrote the paper; A.T. Tan, N. Le Bert, P. L. Lim, B. Young, D.C. Lye and A. Bertoletti reviewed and edited the manuscript; E. S. Lee, E. Chng, P. L. Lim, B. Young and D.C. Lye designed the clinical trial, recruited all the patients and provided the clinical samples; A.T. Tan, N. Le Bert and A. Bertoletti designed and coordinated the study.

## STAR Methods

### Key resource table

2 Key resource table
| Reagent or Resource | Source | Identifier |
| --- | --- | --- |
| **Antibodies** |  |  |
| Anti-human IFN-γ coating antibody | Mabtech | Cat# 3420-3-1000; RRID: AB_907282 |
| Anti-human IFN-γ biotin | Mabtech | Cat# 3420-6-1000; RRID: AB_907272 |
| Brilliant Violet 605 anti-human CD3 Antibody | Biolegend | Cat# 317322; RRID: AB_2561911 |
| Brilliant Violet 650 Mouse anti-human CD4 Antibody | BD Horizon™ | Cat# 563875; RRID: AB_2744425 |
| PE/Cyanine7 Mouse anti-human CD8 Antibody | BD Pharmingen™ | Cat# 557746; RRID: AB_396852 |
| V450 Mouse anti-human CD25 Antibody | BD Horizon™ | Cat# 560355; RRID: AB_1645565 |
| Alexa Fluor® 700 anti-human CD69 Antibody | Biolegend | Cat# 310922; RRID: AB_493775 |
| PE anti-human CD134 (OX40) Antibody | Biolegend | Cat# 350004; RRID: AB_10645478 |
| APC Mouse anti-human CD137 Antibody | BD Pharmingen™ | Cat# 550890; RRID: AB_398477 |
| **Biological samples** |  |  |
| Blood from individuals who received SARS-CoV-2 vaccine | National Centre for Infectious Diseases | N/A |
| **Chemicals, peptides, and recombinant proteins** |  |  |
| Ficoll-paque plus | Cytiva | Cat# 17144003 |
| Zombie NIR Fixable Viability Kit | Biolegend | Cat# 423105 |
| Streptavidin-ALP | Mabtech | Cat# 3310-10-1000 |
| KPL BCIP/NBT Phosphatase substrate | SeraCare | Cat# 5420-0038 |
| 15-mer SARS-CoV-2 overlapping Spike, Nucleoprotein and Membrane peptides | Genscript | N/A |
| **Critical commercial kit** |  |  |
| Ella multianalyte immunoassay | Protein simple | N/A |
| EasySep Human CD4 Positive Selection Kits II | Stemcell Technologies | Cat# 17852 |
| EasySep Human CD8 Positive Selection Kits II | Stemcell Technologies | Cat# 17853 |
| **Deposited data** |  |  |
| No unique dataset was generated | N/A | N/A |
| **Software and algorithms** |  |  |
| Graphpad Prism 9 | Graphpad | https://www.graphpad.com/scientific-software/prism/ |
| Immunospot software | Cellular Technology Limited | http://www.immunospot.com/ImmunoSpot-analyzers-software |
| Kaluza Analysis Software | Beckman Coulter | https://www.beckman.com/flow-cytometry/software/kaluza |

### Resource availability

#### Lead contact

Further information and requests for resources and reagents should be directed to and will be fulfilled by the lead contacts, Anthony T Tan and Antonio Bertoletti.

#### Material availability

This study did not generate new unique reagents.

#### Data and code availability

This study did not generate any unique datasets or new codes.

### Experimental model and subject details

The study design and protocol for the COVID-19 PROTECT study group were assessed by National Healthcare Group (NHG) Domain Specific Review Board (DSRB) and approved under study number 2012/00917. Written informed consent was obtained from all study participants in accordance with the Declaration of Helsinki for Human Research.

3 cohorts of vaccinated healthy individuals were studied (Table 1). Vaccinated individuals were between 22-69 years of age, healthy and with no history of SARS-CoV-2 infection. The first cohort (Inactivated cohort) are healthy individuals who received two doses of BBIBP inactivated vaccine (Sinopharm) given 21 days apart (n=30). The second cohort (Heterologous cohort) are healthy individuals who developed adverse events after receiving a single dose of mRNA vaccine and proceeded to switch to vaccination with two doses of CoronaVac inactivated vaccine (Sinovac) given 21 days apart (n=19). A reference cohort (mRNA cohort) of healthy individuals who received two doses of Spike mRNA vaccine (Pfizer-BioNTech or Moderna) were also analysed (n=79). Blood samples were collected before receiving inactivated vaccines (Inactivated and Heterologous cohort) or mRNA vaccines (mRNA cohort), 21 days and 2-3 months post-vaccination (Figure 1A). Local policy changes recommended the inclusion of a third dose of inactivated SARS-CoV-2 vaccines into the primary vaccination series 3 months after the administration of the second dose. To assess the effects of this booster dose of inactivated vaccines, blood samples of individuals from the inactivated and heterologous cohort were also collected 6 months post-vaccination. A total of 18 individuals in the Inactivated cohort and 7 in the Heterologous cohort were given the booster dose 1-3 months and <1 month before blood collection respectively.

### Method details

#### Peptides

15-mer peptides that overlapped by 10 amino acids spanning the entire protein sequences of Nucleoprotein, Membrane and Spike protein of SARS-CoV-2 were synthesized and pooled into mega-pools. In addition, 55 Spike peptides covering the immunogenic regions of the SARS-CoV-2 spike protein representing 40.5% of the whole Spike protein (SpG peptide pool) were also synthesized, pooled and used to stimulate whole blood as described previously^29^. To measure the T cell response against the Omicron variant, two additional peptide pools were designed for each viral structural protein (Membrane, Nucleoprotein and Spike). The two pools include peptides impacted by the mutations in Omicron, with one consisting of ancestral-derived peptides and another consisting of Omicron-derived peptides covering the same region. Details of all the peptides used are found in Supplementary Tables 1-4.

#### Whole blood cytokine release assay

The whole blood cytokine release assay was performed as described previously^29^. In brief, 320 μl of freshly drawn blood (drawn within 6 hours of venepuncture) were mixed with 80 μl RPMI and stimulated with the indicated SARS-CoV-2 Spike peptide pools (Supplementary Table 1) at 2 μg/ml or with DMSO as a control. After 16 hours of incubation, the culture supernatant (plasma) was collected and stored at −30°C. Cytokine concentrations in the plasma were quantified using an ELLA machine with microfluidic multiplex cartridges that measured IFN-γ and IL-2 according to the manufacturer’s protocol (Protein Simple). The levels of cytokines present in the plasma of DMSO controls were subtracted from the corresponding peptide pool-stimulated samples. The positivity threshold was set at 10x times the lower limit of quantification of each cytokine (IFN-γ = 1.7pg/ml; IL-2 = 5.4pg/ml) after DMSO background subtraction.

#### PBMC isolation

Peripheral blood was collected from all individuals in heparin-containing tubes, and peripheral blood mononuclear cells (PBMCs) from all collected blood samples were isolated by Ficoll-Paque density gradient centrifugation. Isolated PBMCs were either analysed directly or cryopreserved in liquid nitrogen until required.

#### Depletion of CD4+ or CD8+ T cells

CD4+ or CD8+ T cells were positively selected from freshly isolated PBMCs with EasySep Positive Selection Kits II (Stemcell Technologies). After magnetic affinity cell sorting, flow through consisting of CD4-enriched and CD8-enriched PBMCs were collected and used immediately for IFN-γ ELISPOT assay.

#### IFN-γ ELISPOT Assay

The frequency of SARS-CoV-2 specific T cells was quantified as described previously ^29^. Briefly, ELISPOT plates (Millipore Sigma) were coated with human IFN-γ antibody overnight at 4°C. 250,000 to 400,000 total, CD4-enriched or CD8-enriched PBMCs were seeded per well and stimulated for 18 hours with the indicated peptide pools at 1 μg/mL. The plates were then incubated with a human biotinylated IFN-γ detection antibody, followed by streptavidin–alkaline phosphatase (streptavidin-AP) and developed using the KPL BCIP/NBT phosphatase substrate (Seracare Life Sciences). To quantify the peptide-specific responses, spots of the unstimulated wells were subtracted from the peptide-stimulated wells, and the results were expressed as spot-forming cells (SFC) per 10^6^ PBMCs. Results were excluded if negative control wells had more than 30 SFC/10^6^ PBMCs or if positive control wells stimulated with PMA/ionomycin were negative.

#### Activation-induced marker (AIM) assay

Cryopreserved PBMCs were thawed and stimulated with 1 μg/mL mega-pool Spike peptides or equivalent amount of DMSO for 24 hours at 37°C. Cells were then washed in phosphate buffered saline and stained with Zombie NIR Fixable Viability Kit (Biolegend) to exclude dead cells in the analysis. The cells were next washed in FACS buffer with 2mM EDTA and surface markers were stained with the surface markers, anti-CD3, anti-CD4, anti-CD8, anti-CD25, anti-CD69, anti-CD134 (OX40) and anti-CD137 (4-1BB), diluted in FACS buffer for 30 minutes on ice. After two more washes in FACS buffer, cells were resuspended in PBS prior to acquisition with Beckman Coulter CytoFLEX S analyser.

### Quantification and statistical analysis

All statistical analyses were performed using GraphPad Prism, version 9 (GraphPad Software). Where applicable, the statistical tests used and the definition of centre were indicated in the figure legends. Statistical significance was defined as having a P-value of less than 0.05. In all instances, “n” refers to the number of patients analysed.

## Supplemental Information

**Supplementary Figure 1.**
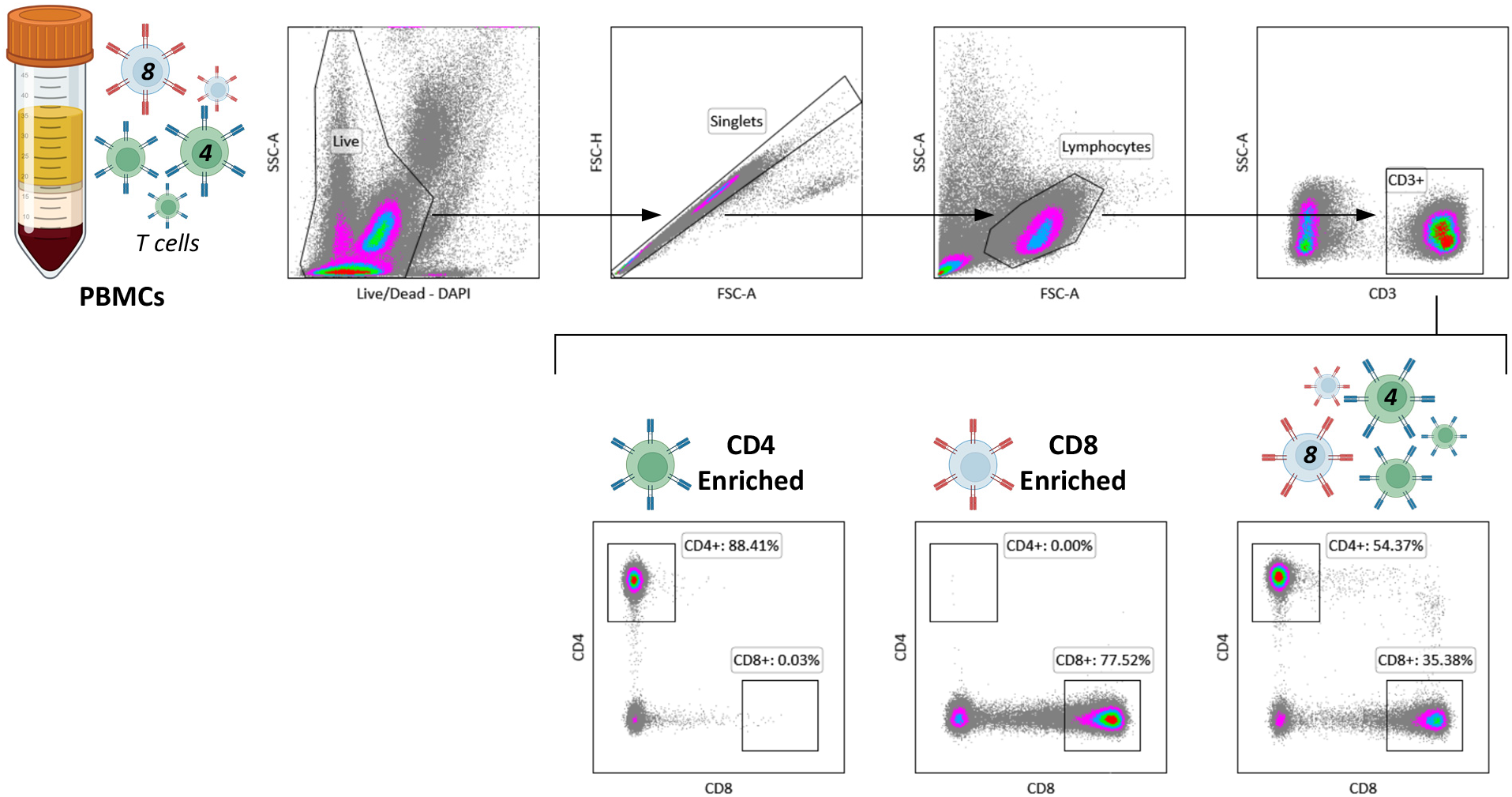
Representative flow cytometry analysis of CD4 enriched, CD8 enriched and total PBMCs.

**Supplementary Figure 2.**
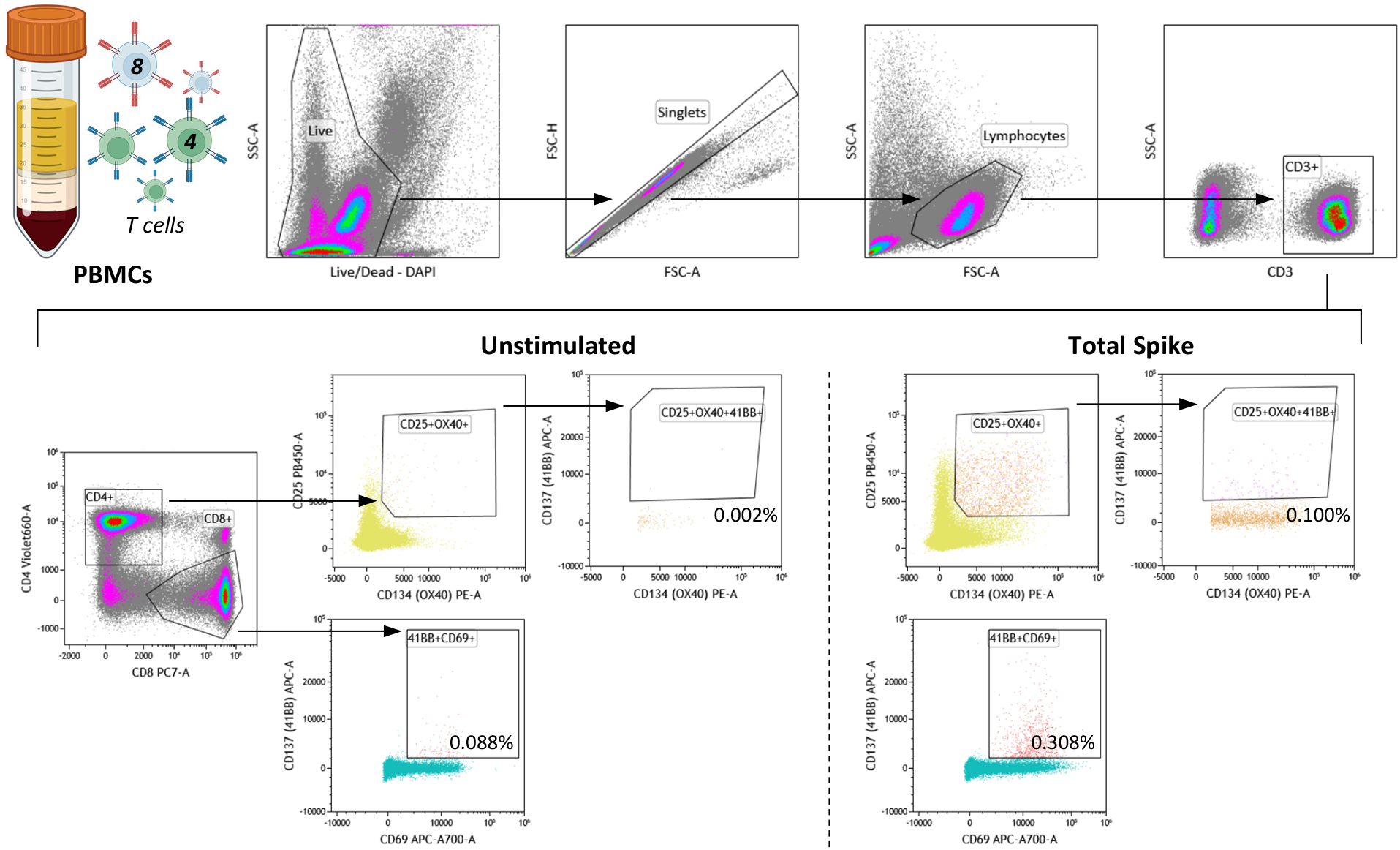
Phenotypic analysis of Spike-specific T cells induced by inactivated SARS-CoV-2 vaccine. PBMCs were isolated from whole blood of individuals in the inactivated cohort 2-3 months post-vaccination. The PBMCs were stimulated with the Spike mega-pool or left unstimulated as a control, and the upregulation of T cell activation markers on the CD4 and CD8 T cell subsets were quantified through flow cytometry. The frequency of AIM+ CD4 (CD25+ OX40+ 41BB+) and CD8 (CD69+ 41BB+) T cells before and after stimulation with Spike mega-pool (n=6) are shown in panel A and B respectively. Differences were analysed with Mann Whitney test and only significant P values are shown (****: P≤0.0001; ***: P≤0.001; **: P≤0.01; *: P≤0.05). Dot plots shows the AIM+ CD4 and CD8 T cell frequencies before and after Spike mega-pool stimulation in a representative individual.

**Supplementary Figure 3.**
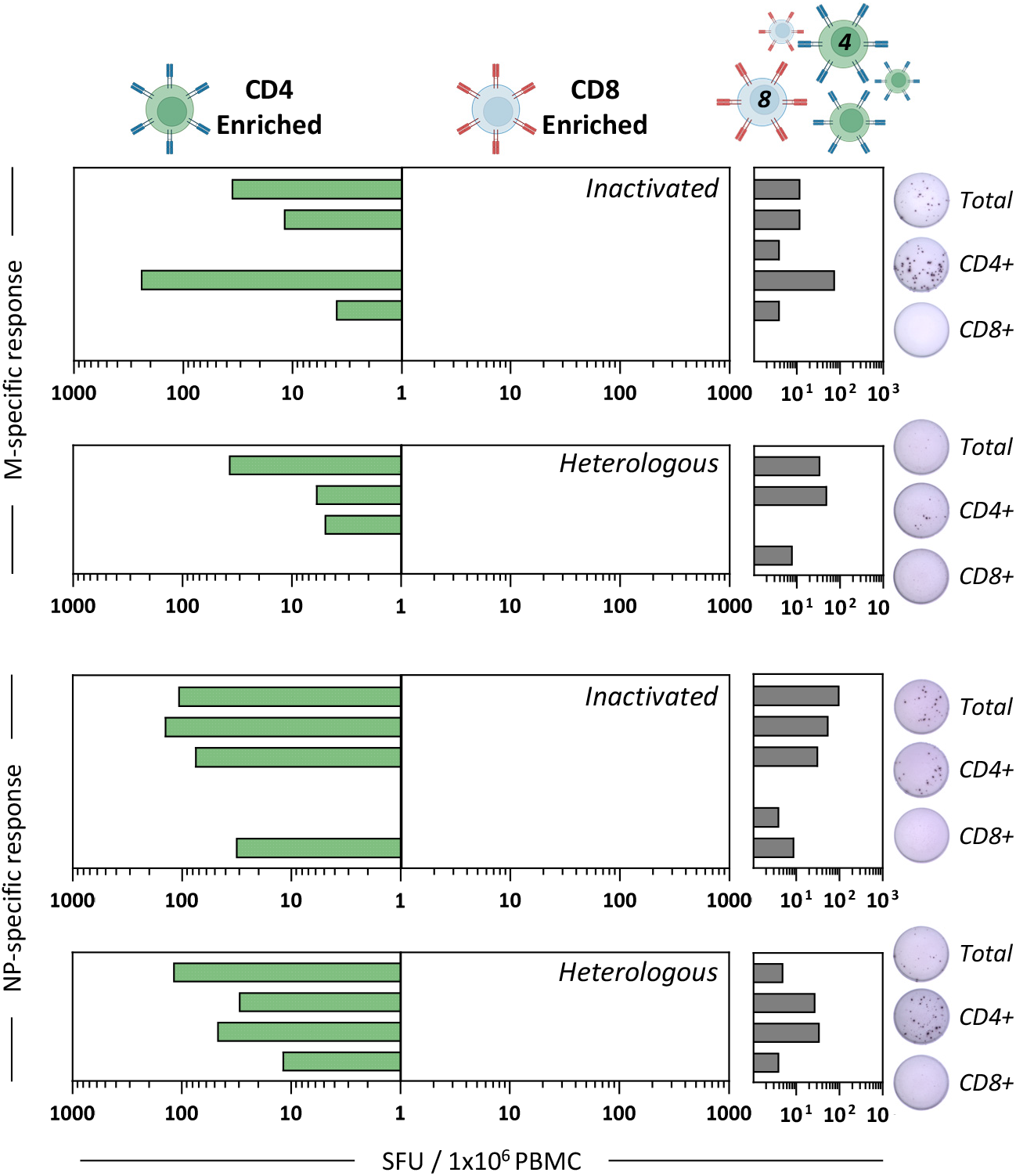
CD4 Membrane- and Nucleoprotein-specific T cells were induced after vaccination with inactivated SARS-CoV-2. PBMCs collected from the 2 cohorts of vaccinees 2-3 months after vaccination (Inactivated: n=6; Heterologous: n=4) were either depleted of CD8 or CD4 T cells through negative selection before stimulation with overlapping peptides covering the entire Membrane and Nucleoprotein for ELISPOT. Total PBMCs were also analysed as a control. Bars denote the IFN-γ SFU quantified for each vaccinee after stimulating CD4-enriched (green), CD8 enriched (blue) or total PBMCs (grey) with Membrane (top) or Nucleoprotein (bottom) overlapping peptides. Representative ELISPOT well images of the respective peptide stimulated cell populations are shown.

**Supplementary Table 1.**
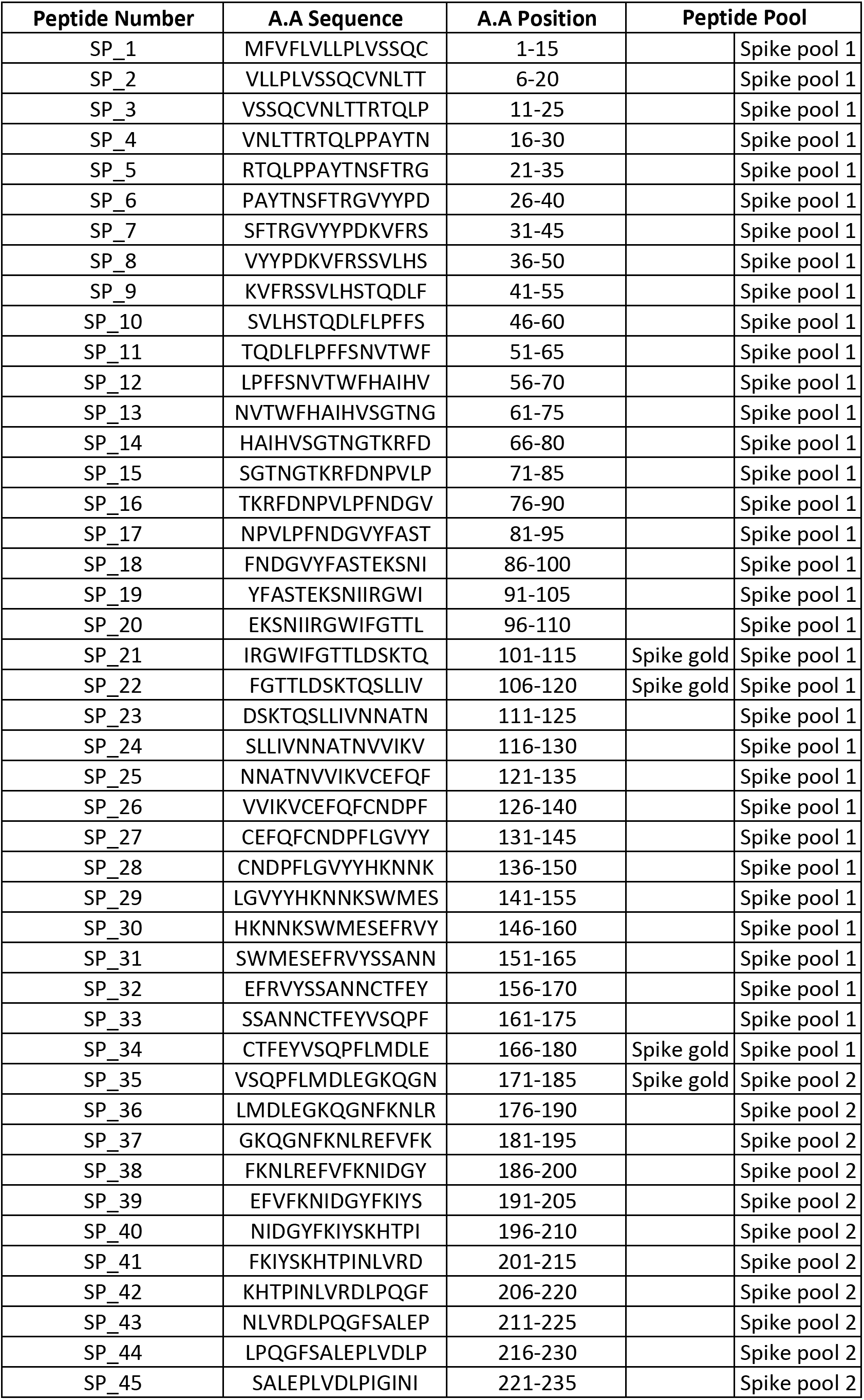
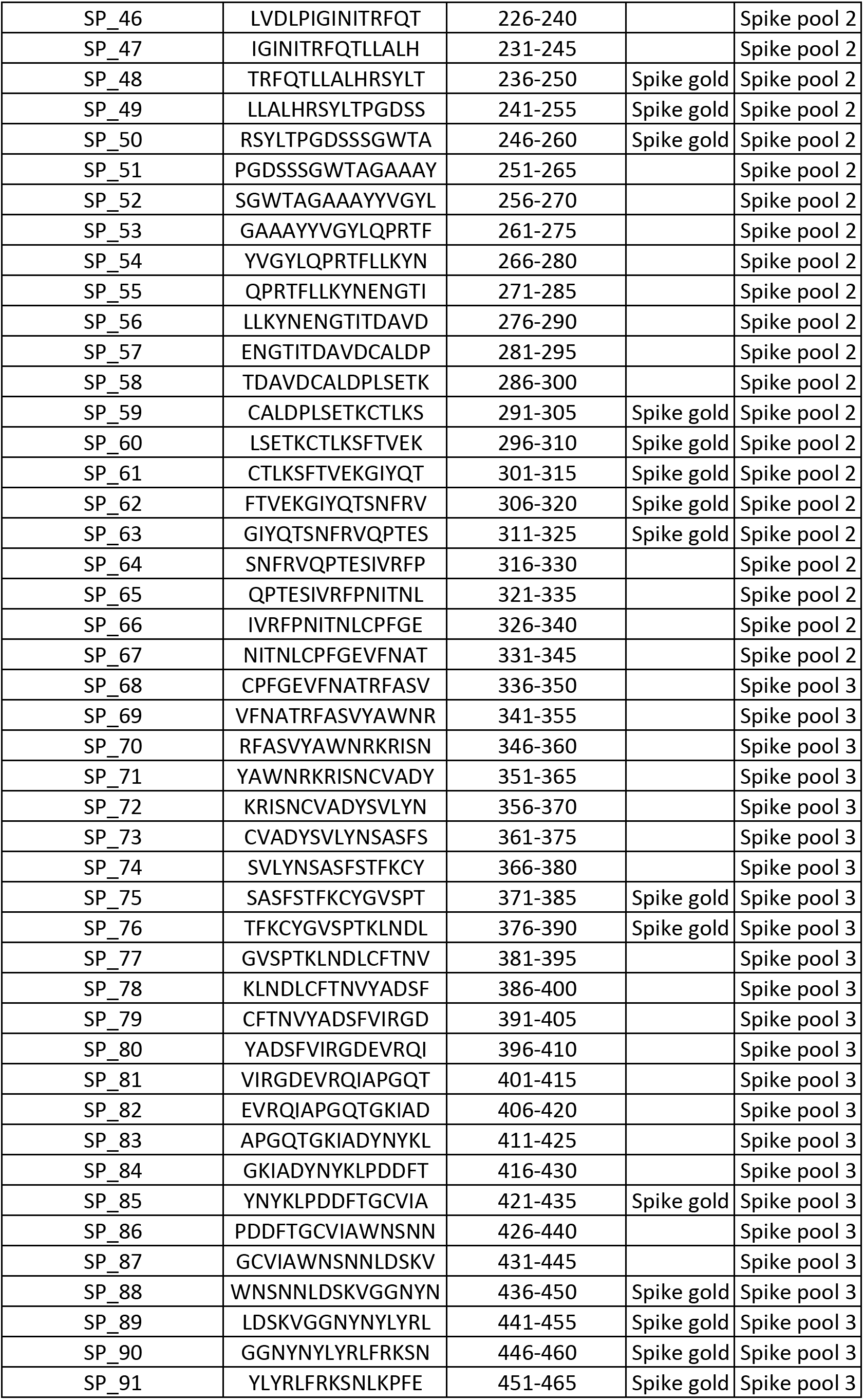
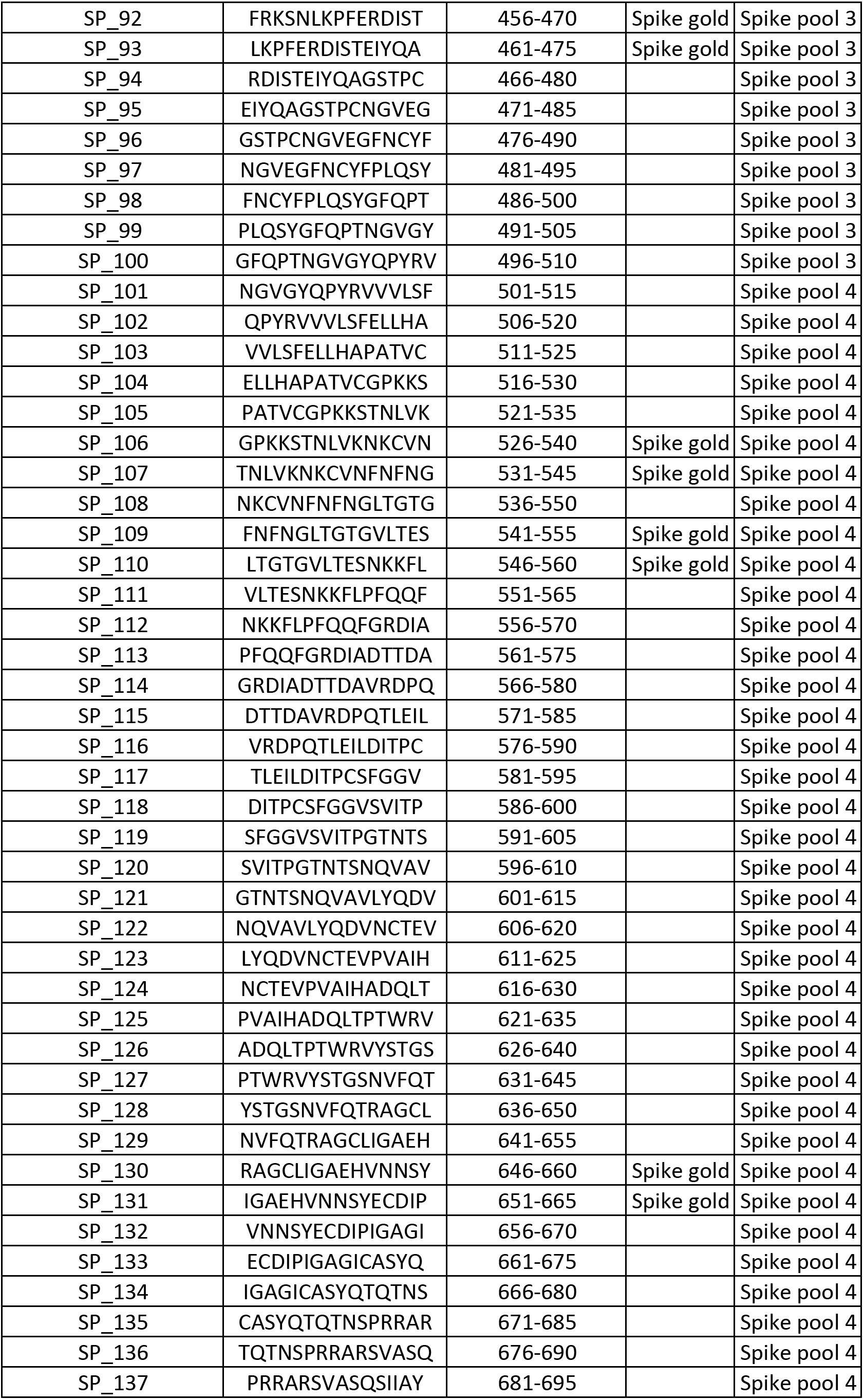
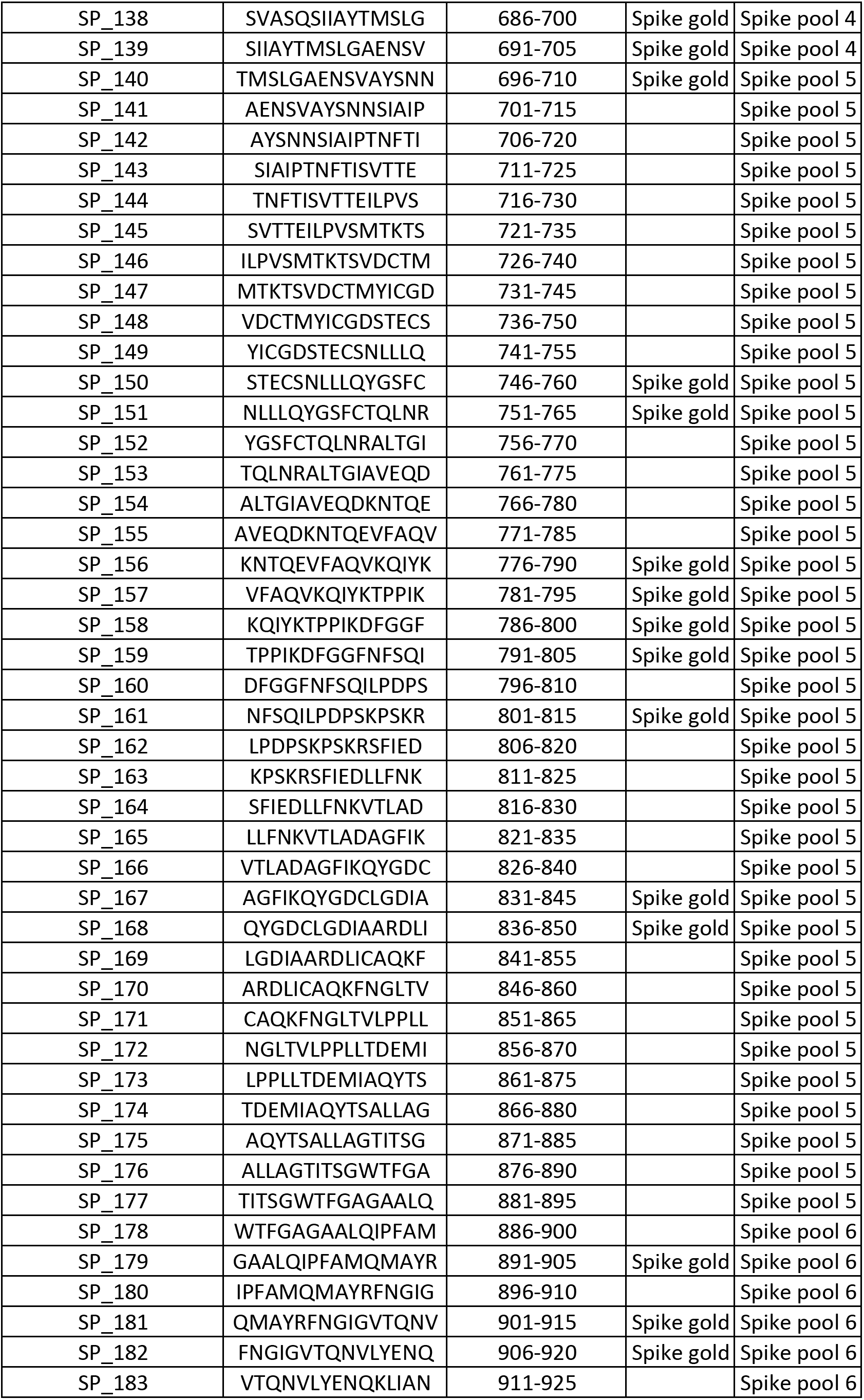
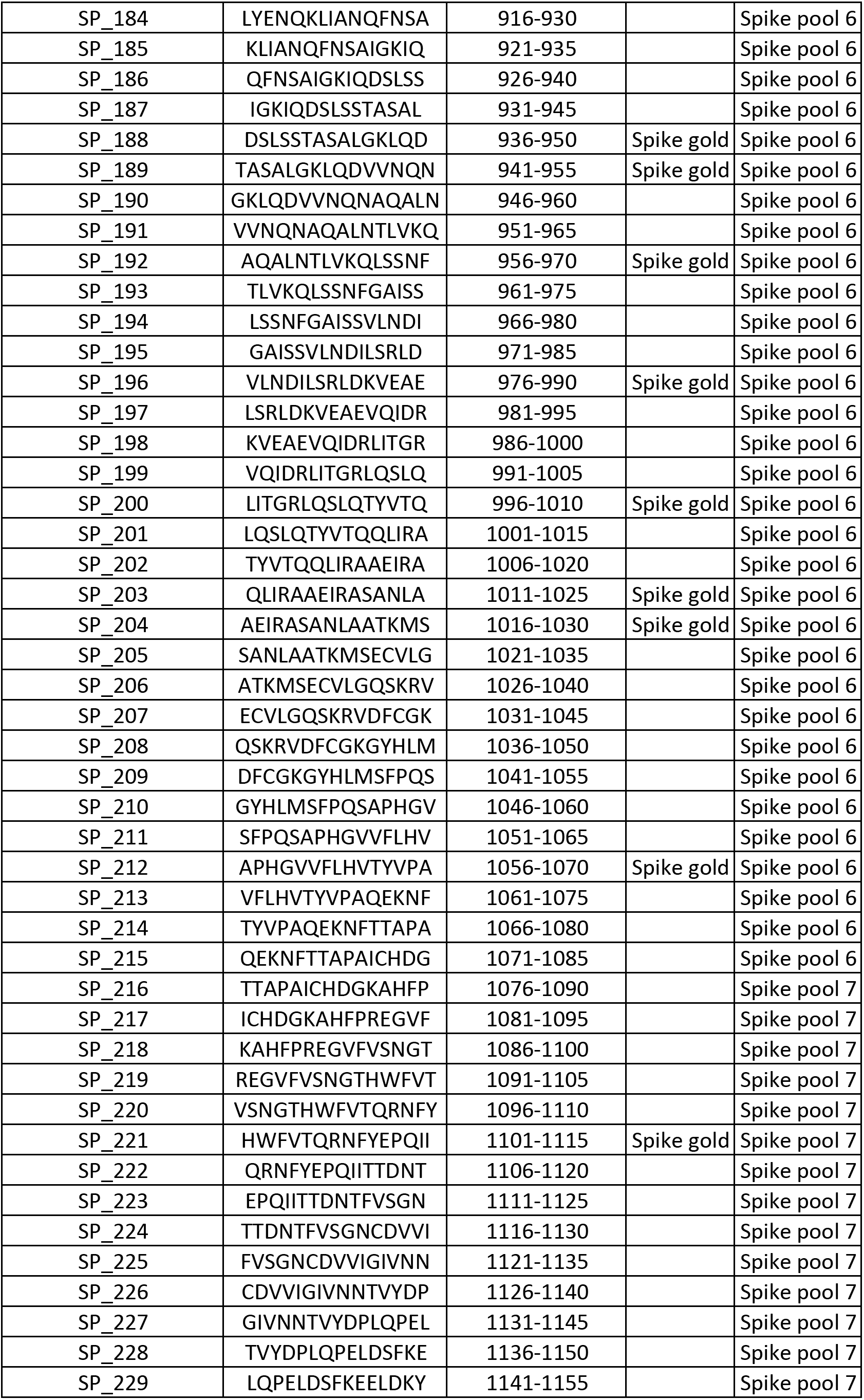
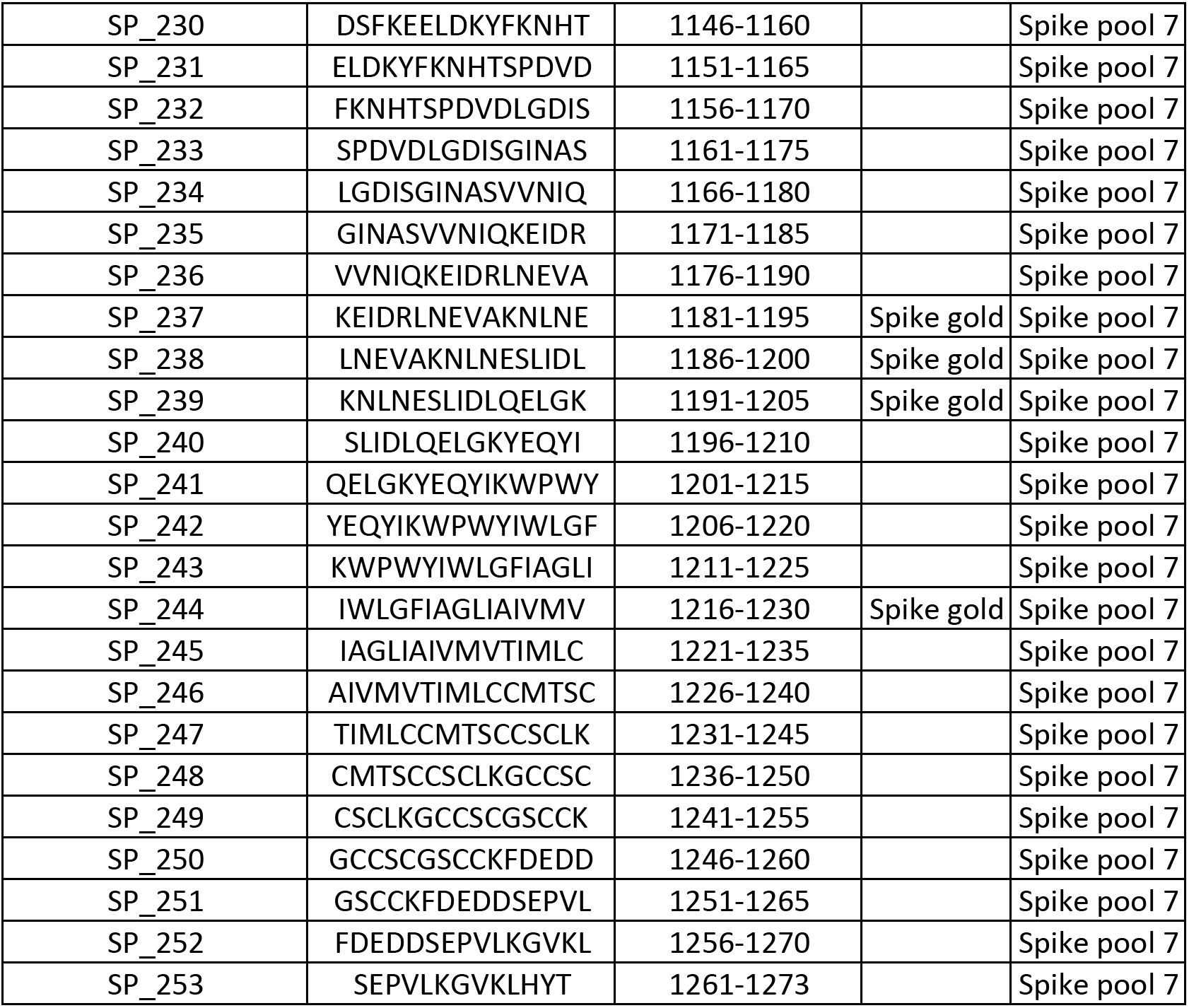
SARS-CoV-2 Spike-specific peptide sequence.

**Supplementary Table 2.**
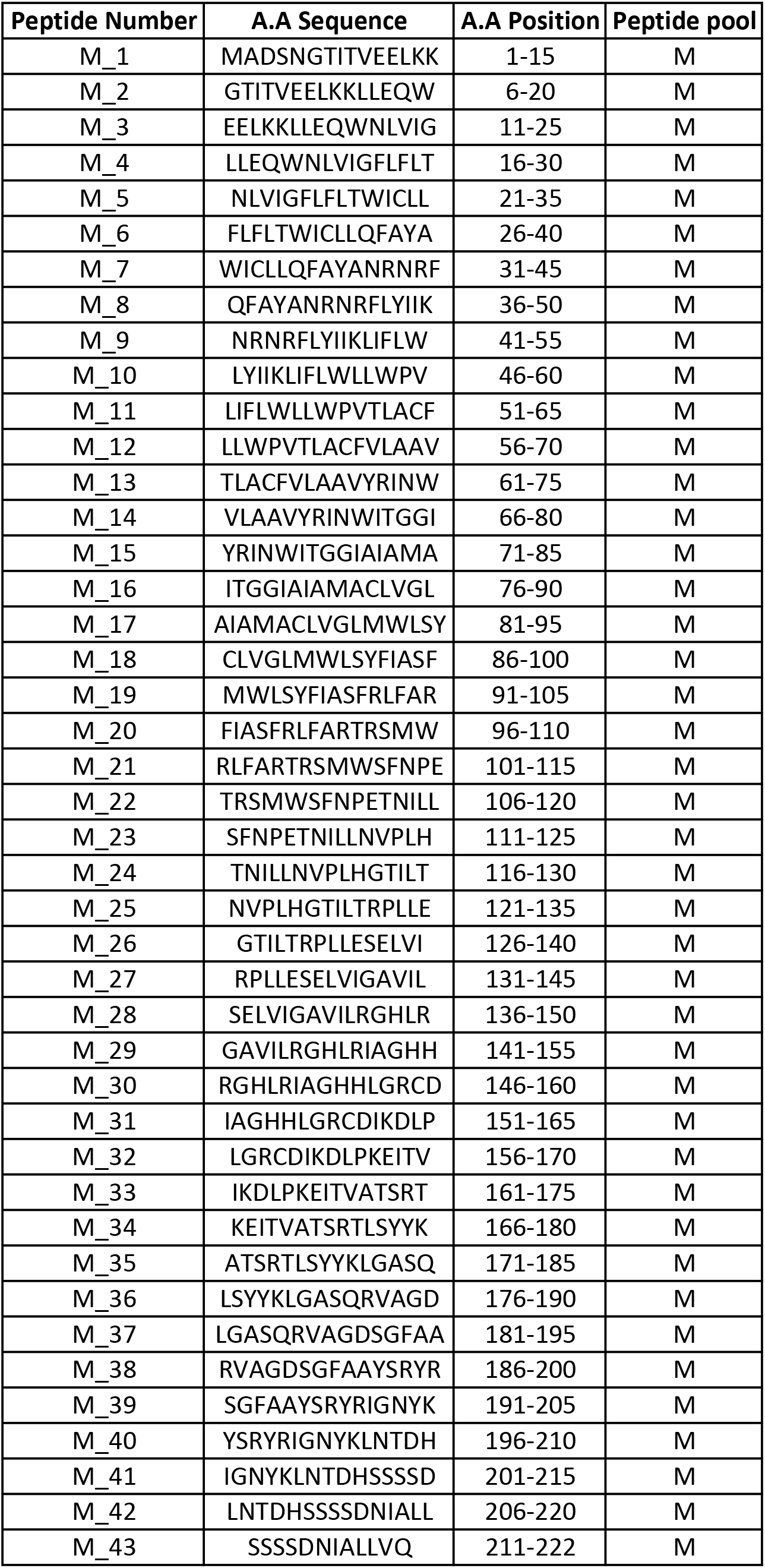
SARS-CoV-2 Membrane-specific peptide sequence.

**Supplementary Table 3.**
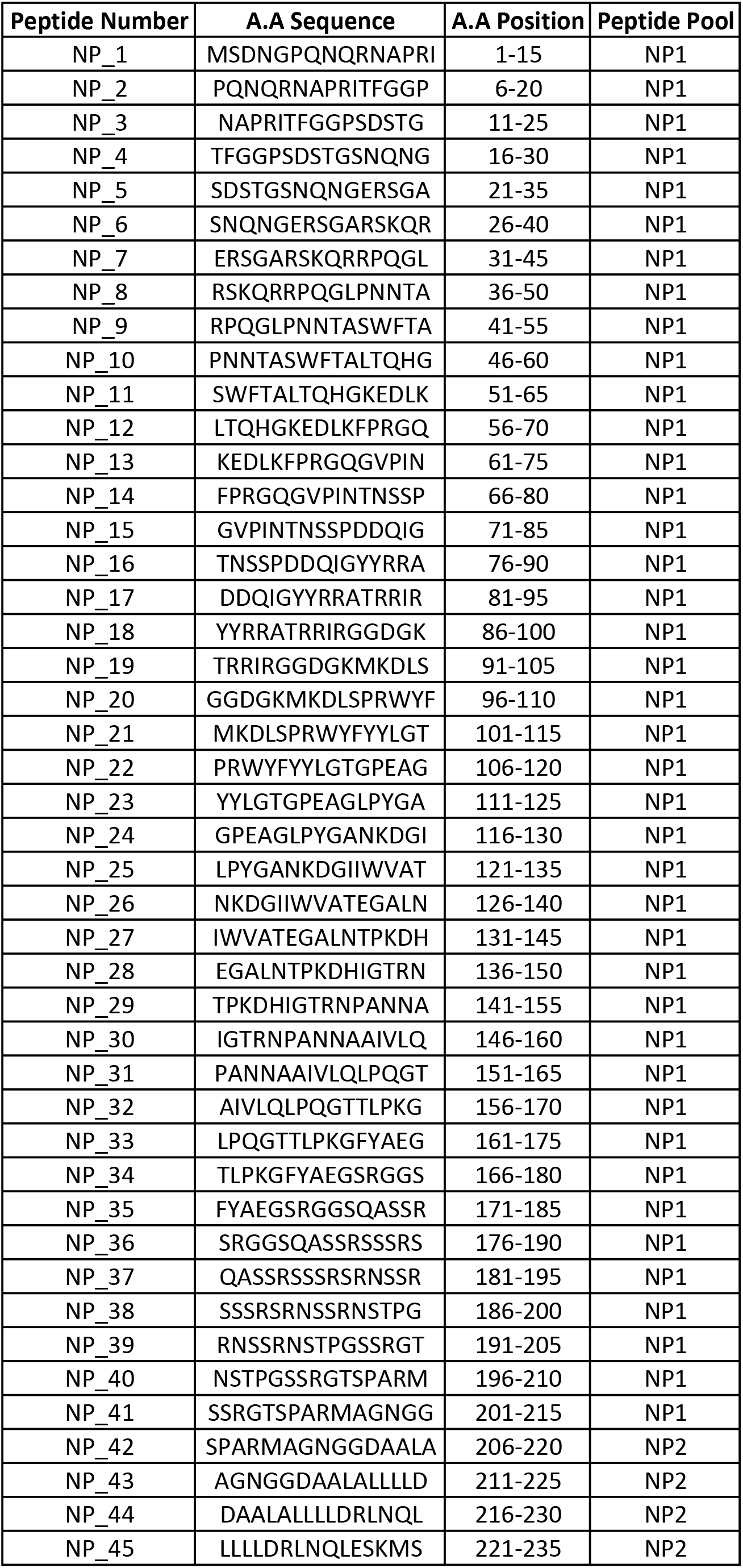
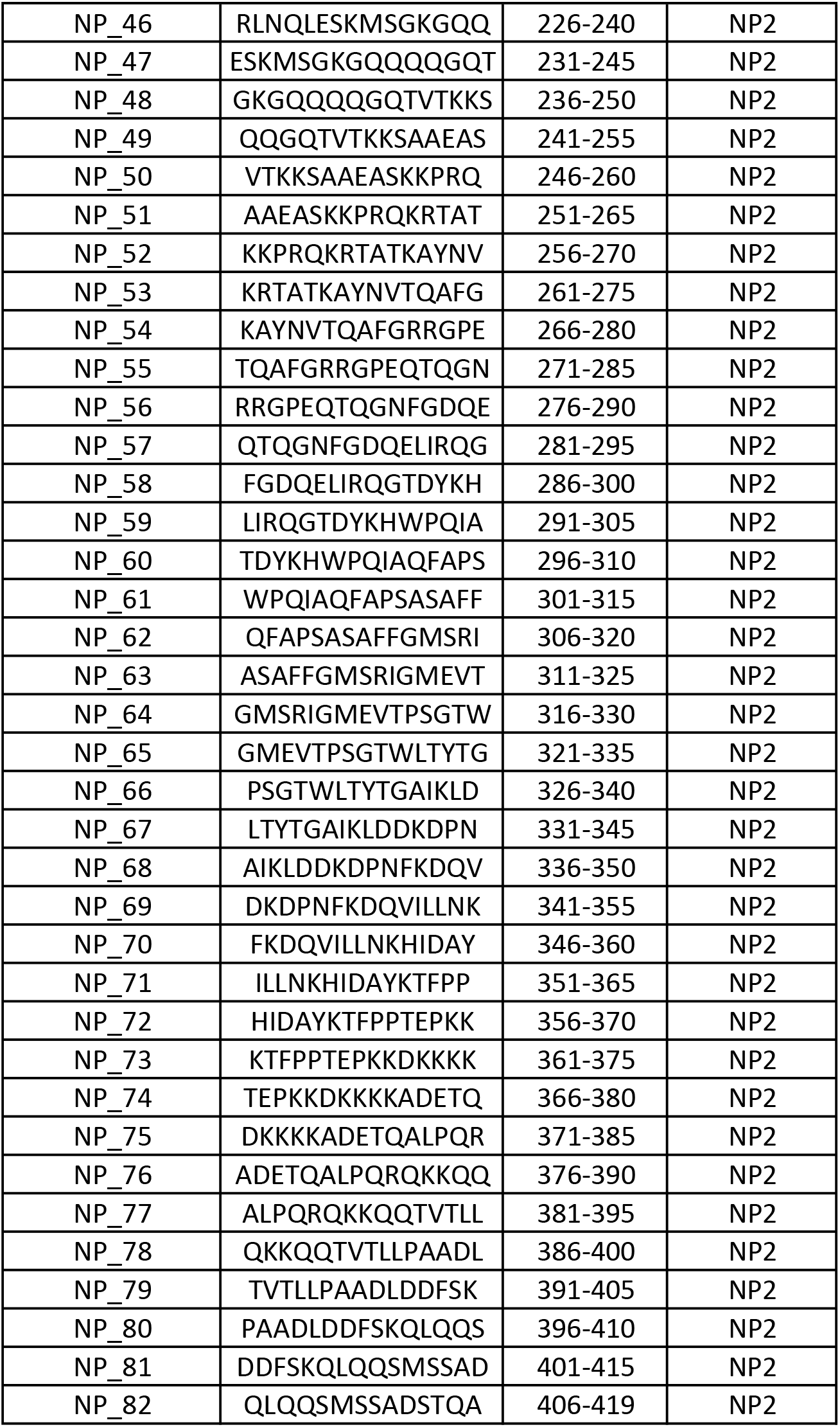
SARS-CoV-2 Nucleoprotein-specific peptide sequence.

**Supplementary Table 4.**
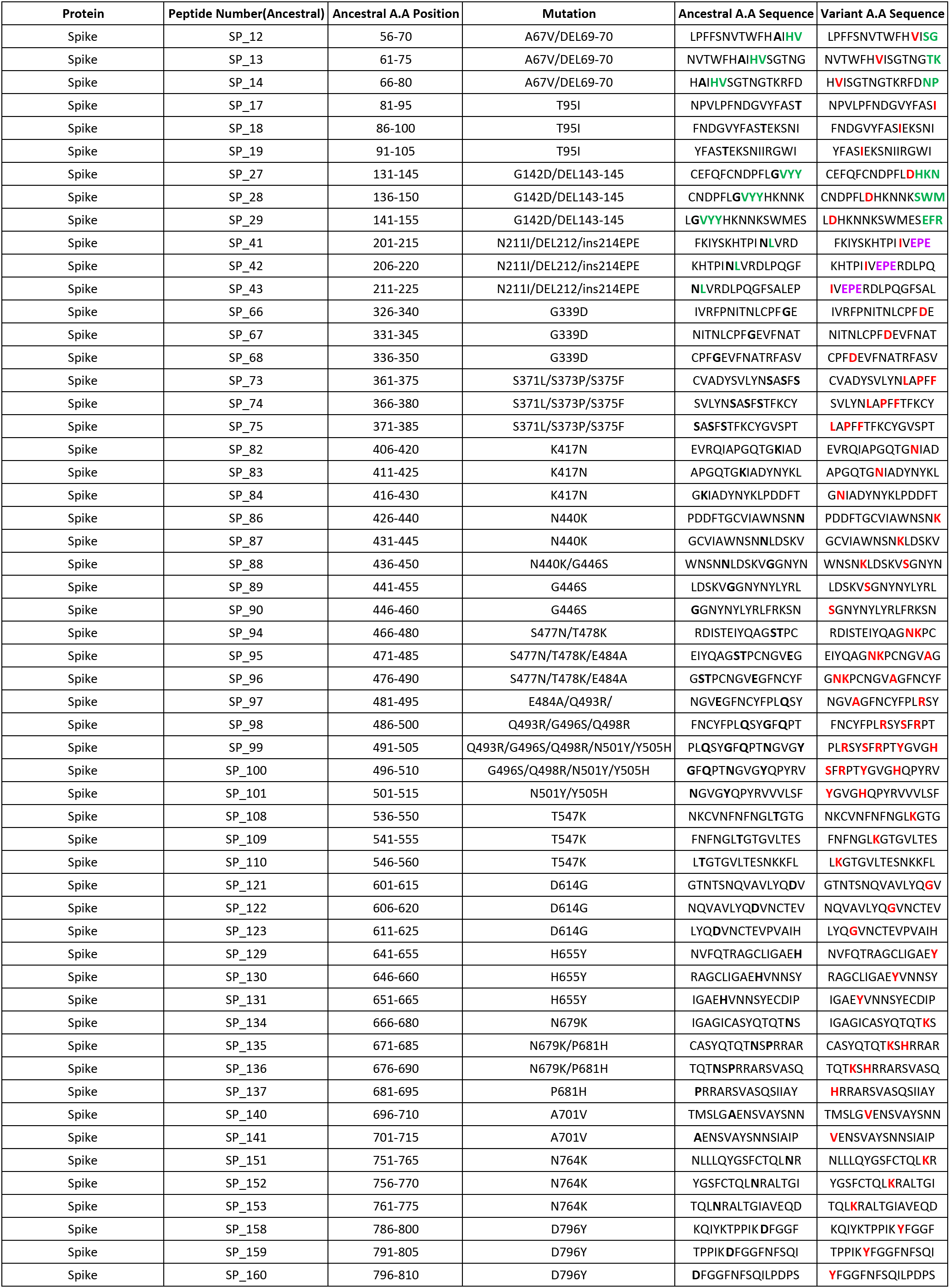
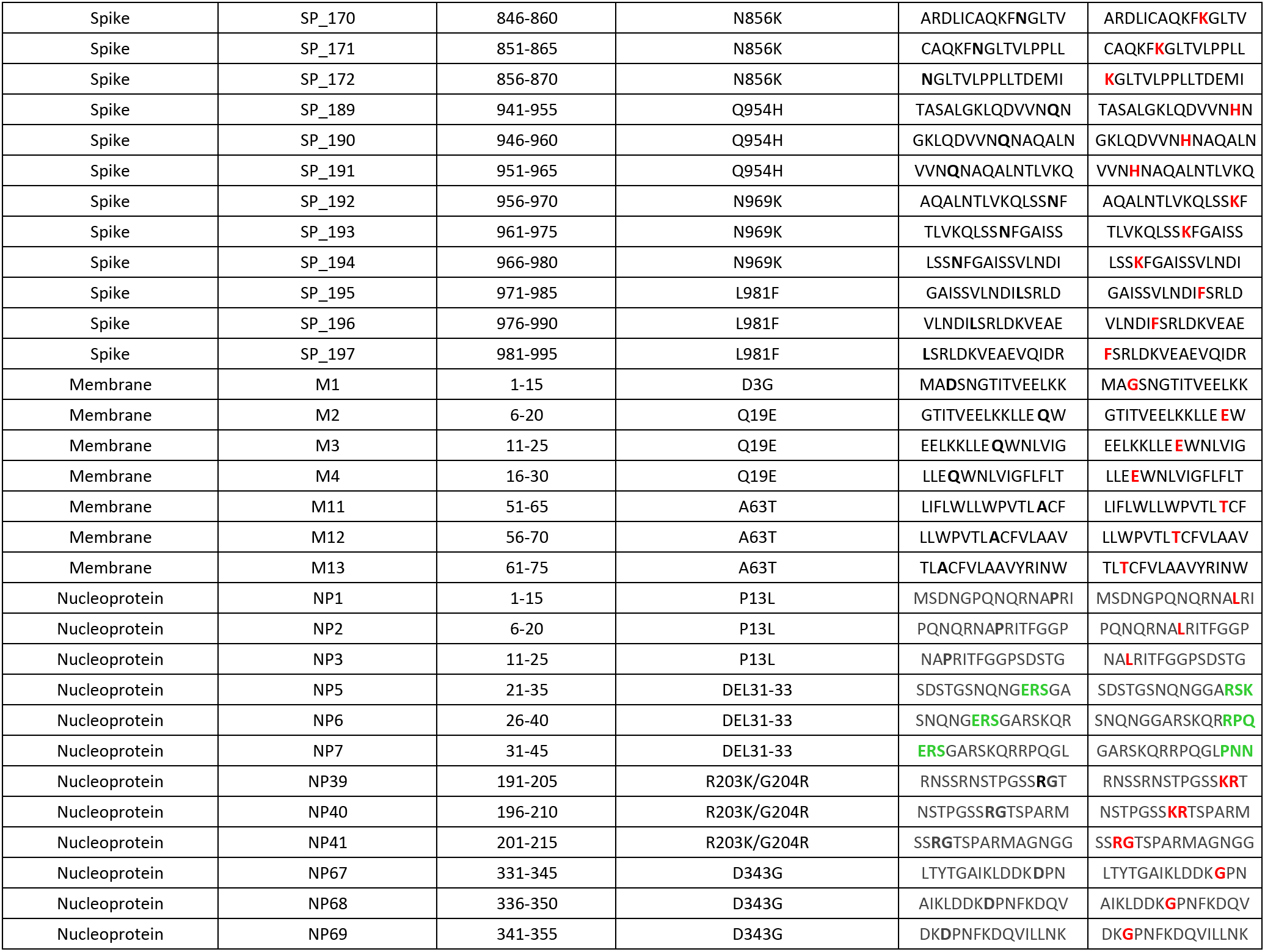
SARS-CoV-2 Omicron VOC peptide sequence.

